# Estimating direct and indirect genetic effects on variation in depressive symptoms in early adolescence: a trio PGS analysis in the MoBa cohort

**DOI:** 10.64898/2026.04.17.26350751

**Authors:** Meseret M. Bazezew, Bernt Glaser, Laura Hegemann, Adrian Dahl Askelund, Jean-Baptiste Pingault, Robyn E. Wootton, Neil M. Davies, Helga Ask, Alexandra Havdahl, Laurie J. Hannigan

## Abstract

**Background:** Early adolescence is a common period of onset for depressive symptoms. In part, this may reflect a developmental manifestation of individuals’ genetic propensities as they undergo physiological and hormonal changes and interact with new environments. Many commonly proposed mechanisms assume direct effects of an individual’s own genes on emerging variation in their depressive symptomatology. However, estimates of genetic influence based on analyses in unrelated individuals capture not only direct genetic effects but also genetic effects from parents and other biologically related family members.

**Aim:** In data from the Norwegian Mother, Father and Child Cohort (MoBa), we used linear mixed models to distinguish developmentally-stable and adolescence-specific direct and parental indirect genetic effects. We examined effects of polygenic scores for major depressive disorder (MDD), ADHD, anxiety disorders, and educational attainment (EA) on depressive symptoms, which were assessed by maternal reports at ages 8 and 14.

**Results:** Children’s own MDD polygenic scores showed adolescence-specific effects on depressive symptoms (βPGS*wave=0.041, [95% CI: 0.017, 0.065]). Developmentally-stable direct effects from children’s polygenic scores for MDD (β=0.016, [0.006, 0.039]), ADHD (β=0.024, [0.008, 0.041]) and EA (β=-0.02, [–0.038, –0.002]) were also evident. The only evidence of indirect genetic effects was a stable effect of maternal EA polygenic scores (β=0.04, [0.024, 0.054]).

**Conclusion:** Direct genetic effects linked to genetic liability to MDD accounted for emerging variation in depressive symptoms in adolescence. These results imply that specific etiological mechanisms related to MDD may become particularly relevant for depressive symptoms during early adolescence compared to at earlier ages.

## 1. Introduction

Depression is a prevalent mental health condition and is projected to become the leading cause of global disease burden by 2030 (WHO, 2012). Adolescence is a sensitive period for the emergence of depressive symptoms and disorders (1–3). The transition from childhood to adolescence is characterised by pronounced biological, social and psychological change, and new potential for exposure to psychosocial stress (1,4–6). During this transitional period, inherited risks, physiological and hormonal development, and potential new sources of psychosocial stress may interact to increase the risk of depression, in part by altering hormonal balance and disrupting neural pathways involved in mood regulation (1,4–9). Given that the increase is greater for girls than for boys (1,10,11), mechanisms may also be sex-specific. Identifying mechanisms with effects specific to this developmental period may inform the development of age-tailored screening, prevention, and intervention strategies.

There is strong evidence that genetic factors play a role in the development of depression (12,13). Some studies suggest that genetic influences may be stronger in early-onset depression compared to adult-onset major depressive disorder (MDD) (8,14–17). Despite the genome being determined at conception, it is possible for genetic factors to become influential only during particular developmental periods. Refining our understanding of the source, timing, and nature of newly-emerging etiological influences on depressive symptoms may help to isolate mechanisms linked to age-related increases in depression (1,7,18,19).

Three previous studies have sought to investigate adolescence-specific genetic influences on depression using polygenic scores (PGS) as indices of specific liabilities (20–22). Two of these have used samples of unrelated individuals. In such designs, associations between children’s genetic liabilities and their depressive symptoms are not straightforward to interpret. This is because studies using unrelated individuals (sometimes called population-based designs) cannot differentiate individual or *direct genetic effects* (those from the child’s own genome on the child’s outcome) and familial or *indirect genetic effects* (those from the parents’ genomes, via the familial or social environment) (23,24). Furthermore, genetic effect estimates in these studies are also at risk of confounding via stratification or assortative mating in prior generations (25).

Within-family designs offer better solution to these challenges, by allowing associations between offspring outcomes and their own genetic liabilities to be separated from both structural confounding and indirect genetic effects operating via parental genetic liabilities for the same traits. Genetic variants influencing depressive symptoms in adolescence via a direct genetic effects pathway may imply mechanisms such as influence on pubertal development or timing, regulation of the release of neurotransmitters in the brain, or control of inflammatory responses to psychosocial stress (26,27). In contrast, genetic effects via an indirect pathway would point to substantially different mechanisms, such as the environment created by parents because of parental genetic liability to various traits, or parents’ capacity to help their child manage stressful situations (28,29).

Using within-family models to look at genetic effects on depressive symptoms during the transition to adolescence, Shakeshaft et al. (22) found more evidence for direct than indirect genetic effects in children’s depressive symptoms. Their results showed that direct genetic effects (transmitted PGS for MDD) predicted higher emotional disorder symptoms between ages 5 and 17, with these direct effects strengthening during adolescence. In contrast, direct effects attributable to ADHD PGS emerged later from age 11 to 17 (22).

In the current study, we used data from the Norwegian Mother, Father, and Child Cohort Study (MoBa), currently the largest available population-based cohort of genotyped child-mother-father-trios, to investigate direct and indirect genetic effects using a within family design on variation in depressive symptoms across the transitional period from childhood to early adolescence. Direct and indirect effects from genetic liabilities for depression and related traits – as indexed by child and parental PGS derived from the results of recent genome-wide association studies (GWAS) – were further separated into those that were stable across the period, and those that were specific to emerging variation in early adolescence.

## 2. Methods

The methods for this study were preregistered and are available at <u>10.17605/OSF.IO/Q2GY6</u>. Deviations from the preregistered plan, which were minor and did not affect any preregistered hypothesis tests, are described and justified in Supplementary Table S1.

### 2.1 Participants and genotyped data

We used data from MoBa (30,31), which is a population-based pregnancy cohort study conducted by the Norwegian Institute of Public Health. Participants were recruited from all over Norway from 1999 to 2008. Women consented to participation in 41% of the pregnancies. The cohort includes 114,500 children, 95,200 mothers and 75,200 fathers. Data on birth year, child sex, parental age at birth and parity were extracted from the Medical Birth Registry of Norway (MBRN), a national health registry containing information about all births in Norway. Statistics Norway (SSB) registry data was also used for information on household income and parental educational attainment. The current study was approved by The Regional Committees for Medical and Health Research Ethics (2016/1702).

The analytic sample comprised a subset of MoBa families with complete genotypic data (32) for mother-father-offspring trios (N = 42,059). The quality control of MoBa genetic data has been described elsewhere (33).

### 2.2 Measures

#### 2.2.1 Outcomes: children and adolescents’ depressive symptoms

Our primary outcome was mothers’ reports of their children’s depressive symptoms on the Short Mood and Feeling Questionnaire (SMFQ) (34)during middle childhood (8 years) and early adolescence (14 years; see Supplementary Method-1). Six maternally reported items consistent across waves were used in the primary analyses to ensure comparability across time. Missingness on these six items (Supplementary Table-S3) was handled using multilevel multiple imputation. After imputation, the SMFQ depressive symptom scores were calculated from the sum of six included items and standardized to have a mean of zero and a standard deviation of 1.

In cross-sectional sensitivity analyses, we additionally analysed the full 13-item version of the mother-reported SMFQ at age 8 and children’s own reports of their depressive symptoms, which were available only at age 14 (13 items).

#### 2.2.2 Exposures: polygenic scores

We used PGS as exposures indexing genetic liability for depression and related traits. We constructed PGS using summary statistics from the recent Psychiatric Genomics Consortium GWAS of major depressive disorder (35). We additionally included PGS for 3 other traits that have been demonstrated to explain variation in childhood emotional problems elsewhere (36–38), namely: ADHD (39), anxiety (40) and educational attainment. We computed PGS using LDpred2 (41), with the option ‘LDPred2-auto’ (for full details, see Supplementary Methods 1).

#### 2.2.3 Covariates

We selected covariates that represent potential confounders of the relationship between measured genetic liabilities and emerging adolescent depressive symptoms (Supplementary Method-1 and Table-S2). Typically, these are factors (i.e. household income, parental education, child sex, Parity, parental age at birth) that can bias effects estimated in GWAS studies, and which may also affect depressive symptoms. These covariates are included directly into the regression model. Where possible, we sought to avoid including potential mediators of the PGS-depressive symptoms associations (e.g., measures of parental mental health) as covariates to avoid overadjustment bias (see Discussion section for an overview of this issue).

### 2.3 Data Analysis

#### 2.3.1 Missing data

We restricted the analytic sample to individuals with complete mother-father-child trios with genotype data. Within this sample, we set out to impute missing phenotypic information. Consequently, exposures (PGS) were complete, and missingness was mostly in outcomes and covariates. We investigated the extent of missingness and the missingness mechanism before deciding on the missing-data handling strategy (Supplementary Method-1 and 2, Table S3-S6).

Based on various missing mechanism test results, we assumed that data were Missing at Random (MAR). Therefore, we performed multilevel multiple imputation (MMI) to impute item level and covariate missingness using multiple imputation by chained equations (MICE) using the *mice* package v3.16.0; (42) in R Statistical Software (v4.4.1; R Core Team 2024) (43). Full details of the specification of the MMI models are available in (Supplementary Method-3).

Sensitivity analyses using complete case data (CCA) were conducted to highlight any impact, beyond the expected increase in precision, of the multiple imputation on results.

#### 2.3.2 Statistical analysis

We aimed to estimate the associations of children’s genetic liabilities – as well as those of their parents – on depressive symptoms. In particular, we were interested in isolating effects on variation in symptoms emerging in early adolescence (adolescence-specific effects).

First, for depressive symptoms measured at 8 and 14 years respectively, we ran the following models independently: a) bivariate linear mixed models, in which with only a single family member’s PGS for a given trait included as a predictor in a given model; and b) linear mixed models, in which all family members’ PGS for a given trait were included as predictors; c) fully adjusted linear mixed models, in which covariates were added to the models described in (b) above. In these models, random effects were included to account for within-family (across sibling) variation.

Next, we extended the models to predict variation in outcomes from both waves simultaneously, adding a random effect for within-individual (across time) variation, and additionally estimating interaction effects between PGS for each member of the mother-father-child trio, and an indicator for wave of measurement (0/1 coded for the 8-year and 14-year waves respectively). In these models, we were therefore able to estimate both stable (across both timepoints) and adolescence-specific (variation specific to the adolescent timepoint) direct and indirect genetic effects on depressive symptoms. Full specifications of all analytic models are available in Supplementary Method-4.

False discovery rate (FDR) was applied to control for multiple testing (44), adjusting across 4 tests for 4 different exposure PGS within effects from each family member (see preregistration for pre-specified hypothesis tests). All models with exposure-by-wave interactions also included covariate-by-wave interaction terms to account for time-varying confounding.

### 2.4 Availability of data and code

The consent obtained from MoBa participants does not allow for the sharing of individual level data with analyses. Data from MoBa, MBRN and SSB used in this study can be made available to researchers, provided approval from the Norwegian regional committees for medical and health research ethics (REK), compliance with the EU General Data Protection Regulation (GDPR) and approval from the data owners.

All analyses were conducted in R version 4.4.1. Phenotype data were processed prior to analysis from MoBa and MBRN using the *phenotools* R package version *0.4.2* (45). Relevant preparatory and analytic code for the project is available at https://github.com/psychgen/trioPGS-adolescent-depression.

## 3. Results

A total of 42,059 families with complete trios available for PGS were included. Prior to imputation, complete data on depressive symptoms score (via the SMFQ) were available for 44.1% of the sample at age 8 and 27.6% at age 14. The proportion of missingness on covariates was less than 2%. The proportion of male to female children in the study was similar. The average depressive score before imputation was 6.9 (SD=1.29) at age 8 and 7.0 (SD=1.85) at age 14. The genotyped analytic sample and broader MoBa sample were broadly similar across key characteristics (see Supplementary Table-S4), but we observed some evidence of selective attrition (Supplementary Table S7 and S8). All subsequent results, except the CCA sensitivity analyses, are based on analyses of multiply imputed data.

## Direct and indirect genetic effects on depressive symptoms at age 8

In bivariate models of depressive symptoms measured at age 8, child genetic liabilities to MDD and ADHD, as measured by PGS, were associated with their own depressive symptoms (β=0.032, 95% CI: 0.021-0.043, *pFDR* < 0.001) and (β=0.031, 95% CI: 0.019-0.043, *pFDR* < 0.001) respectively. In trio models, after adjusting for parental PGS and other covariates, child genetic liabilities to MDD and ADHD remained significantly associated, but effects were attenuated to (β=0.023, 95% CI: 0.006–0.04, *pFDR* = 0.018) and (β=0.024, 95% CI: 0.008–0.041, *pFDR* = 0.016) respectively (supplementary Table-S11 and S12). Significant effects from mothers’ and fathers’ genetic liabilities in bivariate models (Figure 1 and supplementary Table S11 became non-significant in the fully-adjusted trio models, with the exception of mothers’ genetic liability to EA which was positively predictive of depressive symptoms at age 8 (β = 0.04, 95% CI: 0.024,0.055, *p* < 0.001; Figure 1 and supplementary Table S12). Results from trio models without adjustment for covariates are provided in Supplementary Table S9.

**Figure 1:**
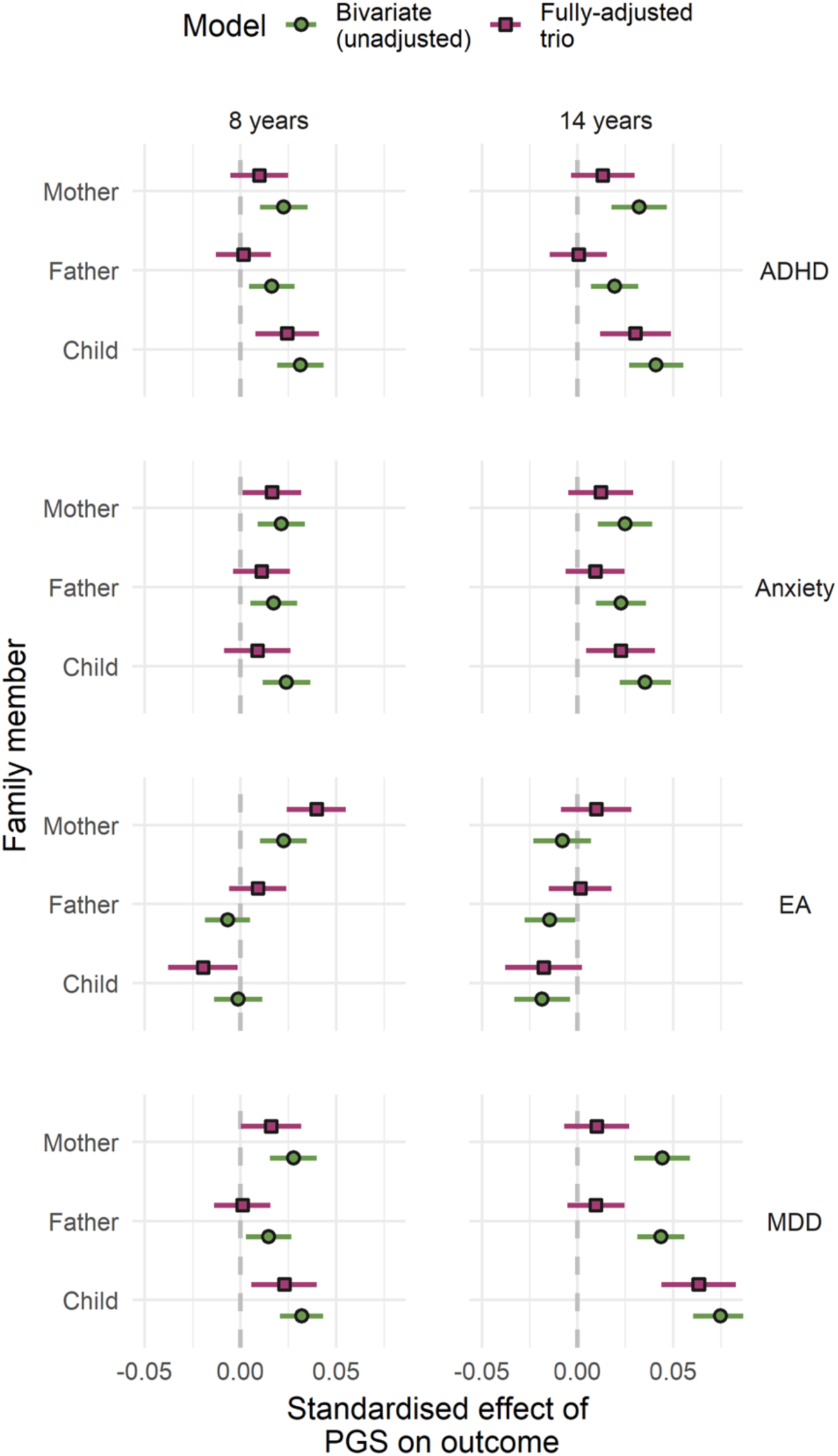
Associations between genetic liabilities and children’s depressive symptoms at age 8 and 14 with and without mutual adjustment for other family members’ PGS and measured confounders **Note:** MDD-major depressive disorder, ADHD-Attention Deficient/Hyperactivity disorder, EA-Educational attainment.

## Direct and indirect genetic effects on depressive symptoms at age 14

In bivariate models, all child genetic liabilities (MDD, ADHD, Anxiety and EA) showed an association with depressive symptoms at age 14. After adjusting for all family members’ genetic liabilities and covariates, the results remained quite consistent, except genetic liability to EA became weakly associated (β=0.045, 95% C.I: –0.038, 0.002, *p*FDR=0.083) (Figure 1, and supplementary Table S11). Both parents’ genetic liabilities to all exposures (except maternal genetic liability to EA) were initially associated with depressive symptoms at age 14 in bivariate models. As with similar estimates at age 8, all parental effects become statistically non-significant in a fully adjusted trio model (Figure 1, and supplementary S12). See supplementary Table-S10, for results from trio models without adjustment for covariates.

## Direct and indirect genetic effects on variation in symptoms of depression specific to early adolescence

Our results indicated that only direct genetic effects from MDD genetic liability showed specificity depressive symptoms in early adolescence. A one SD increase in children’s MDD PGS increases depressive symptoms by 4.1% of a standard deviation *more* at age 14 compared to at age 8 (βPGS*wave=0.041, 95% CI: 0.017, 0.065, *p=*0.004) (Supplementary Table-S13). Children’s genetic liabilities to MDD (β=0.016, 95% CI: 0.0058, 0.039, *p=*0.018), ADHD (β=0.024, 95% CI: 0.008, 0.041, *p=*0.016) and EA (β=-0.02, 95% CI: –0.038, –0.002, *p=*0.042) showed stable association with depressive symptoms (i.e., across both waves), but there was little evidence that direct genetic effects other than MDD specifically differed in early adolescence compared to childhood. The *indirect* genetic effect of maternal genetic liability to EA was predominantly stable across both age 8 and 14 (β=0.04, 95% CI: 0.024, 0.055, *p*<0.001) Figure 2. While this effect decreased somewhat at age 14 relative to age 8 and the shift was not significant (βPGS*wave = –0.03, 95% CI: – 0.053, –0.007, *p*=0.05). Sex stratified estimates are presented in supplementary Fig S4-Fig S6.

**Fig 2.**
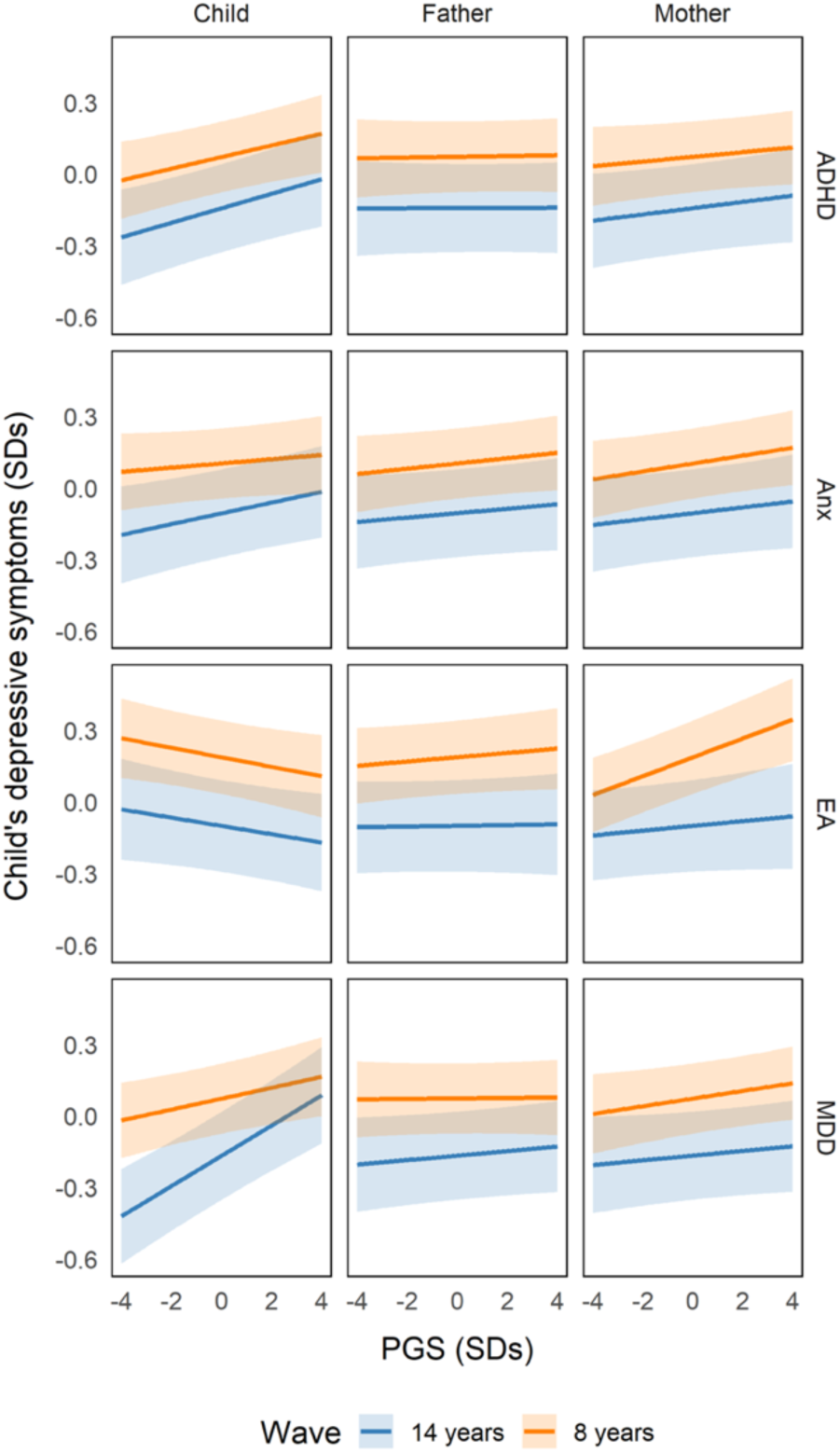
Association of genetic liabilities and emerging variation of depressive symptoms between mid-childhood and early-adolescence.

## Sensitivity analyses

### Complete Case Analysis (CCA)

In CCA, we observed consistent results with those based on multiply imputed data for direct genetic effects (Supplementary Table S13). For indirect genetic effects, maternal genetic liabilities for MDD and anxiety additionally predicted depressive symptoms at age 8 in CCA, and paternal genetic liabilities for MDD and anxiety were associated with children’s depressive symptoms at age 14. In the final model incorporating outcomes across both waves, maternal indirect genetic effects for MDD and anxiety were the only effects across waves that were not present in the primary analyses (Supplementary Result 2, Table S14-S16). A comparison of CCA and MMI results is provided in Supplementary Result 3.

### Additional sensitivity analyses

Results using mother reported full 13-item scale at age 8 on an imputed data, and child-reported items at age 14 using CCA (since the MMI model failed to converge) are provided in Supplementary Table S17 and S18.

## 4. Discussion

We examined whether direct and indirect effects from genetic liabilities for major depressive disorder, ADHD, anxiety, and EA contributed variation in depressive symptoms specific to early adolescence. We found that children’s PGS for MDD, ADHD, and EA were stably associated with depressive symptoms across middle childhood and early adolescence. We also saw evidence of adolescence-specific direct effects from genetic liabilities to MDD. We found limited evidence for parental genetic effects, except for maternal EA polygenic scores, which showed an association with depressive symptoms that was consistent across middle childhood and early adolescence.

### 4.1 Direct genetic effects on depressive symptoms between childhood and early adolescence

In longitudinal models assessing effects of genetic liabilities on depressive symptoms across middle childhood and early adolescence, children’s MDD PGS showed both a stable and adolescence-specific effect. This may indicate that this measure of MDD genetic liability captures variation linked to direct mechanisms involved in adolescent depression. This aligns with previous longitudinal findings (21,22), However, it is important to note that the MDD GWAS underlying the PGS was primarily conducted in adult samples. Consequently, it is possible that associations with depressive symptoms increased in adolescence because participants are approaching the age range of the population from which the PGS was derived, thereby increasing its predictive validity (46) – rather than due to adolescence-specific effects. Nonetheless, evidence that direct effects of MDD genetic liability becomes stronger across the transition period might have additional important implications. Direct effects are likely a result of mechanisms that involve changes in the individual rather than their environmental or social context *per se*. Such mechanisms may still include developmental shifts in exposure or responsivity to environments (47), but these are likely to be attributable – in most part – to genetically influenced characteristics in the individuals themselves.

Although children’s anxiety PGS was associated with higher depressive symptoms at age 14 but not age 8 in cross-sectional models, the shift was not significant in our longitudinal models. Nonetheless, our estimates are broadly consistent with previous reports of increasing effect sizes and variance explained for genetic liability to anxiety between ages 5 and 17 (22), and similar trajectories between age 11 and 18 (21). This points to adolescence as a period when the associations between genetic liability to anxiety and depressive symptoms strengthen.

In contrast, in a longitudinal model, our results for ADHD and EA PGS seem to indicate that these are largely stable sources of direct genetic effects across development. Future research with follow-up longer into adolescence would be necessary to confirm these patterns.

### 4.2 Indirect genetic effects on depressive symptoms between childhood and early adolescence

We found little evidence of indirect genetic effects from parental genetic liabilities for MDD, ADHD, or anxiety on depressive symptoms at either age 8 or 14. This is consistent with prior literature (22). These results suggest that parental genetic liabilities to MDD, ADHD, and anxiety are unlikely to affect offspring depressive symptoms via indirect effects. Conversely, maternal EA PGS was found to be consistently positively associated with child depressive symptoms across the transition from childhood and adolescence – though the effect appeared to weaken substantially at age 14. There are several possible explanations for this result. First, it may reflect genuine exposure effects of genetically influenced maternal behaviour on depressive symptoms across middle childhood and adolescence. Second, the association may have resulted from mothers with a higher genetic liability to EA having an increased tendency to observe and report depressive symptoms among their children. Third, because indirect genetic effect estimates from trio PGS models remain vulnerable to structural confounding (48), potentially including the impact of assortative mating and population structure (25), it may be that these effects reflect residual confounding. Such confounding has been shown, in other within-family designs, to be ‘baked-in’ to the results of EA GWAS (49,50).

#### Limitations

There are several important limitations to consider when interpreting these findings. First, interpretation of our results should also include consideration of potential biases related to selection and attrition within MoBa, especially concerning factors relevant to the traits examined in this study such as the cohort’s overrepresentation of families with higher socioeconomic status (51). Second, indirect genetic effect estimates, especially from PGS calculated from population based GWAS, are known to capture not only true “genetic nurture” (the influence of parents’ genes on their child’s outcomes through the environment they provide, rather than through genes the child directly inherits) but also residual confounding from assortative mating, population structure, related individuals (e.g., siblings or multigenerational influences), and unmeasured environmental factors unrelated to parental genotype, such as neighbourhood disadvantage or life events can further complicate interpretation (25,48,52). Third, maternal reporting bias (e.g., the depression-distortion effect) (53), although the overall bias may not be large (54), possibly inflating associations between maternal genetic risk and child outcomes. Multiple informants or objective measures would help mitigate this bias (53,54). Fourth, by refining genomic discovery based on age-specific associations, as has been done for other conditions (e.g., autism) (16,55), depression-related genetic instruments may be further tailored for use in understanding mechanisms in the future. Fifth, although we sought to exclude mediators from our list of model covariates to minimize overadjustment bias, some included covariates (e.g., income and education) may have been both confounders and mediators. Although their inclusion risks biasing down our estimates, we adjudged that controlling their potential confounding bias – which has the potential produce spurious effects – was the most conservative option. Finally, because analyses were restricted to participants of European ancestry, the generalizability of these findings to other populations is limited.

## Conclusions

By investigating the direct and indirect genetic effects on depressive symptoms from middle childhood to early adolescence, we partitioned specific sources of genetic liability in terms of when – and via whom – they exert their effect. By refining genetic instruments according to when, across development, associations between genetic variants and traits arise or peak, and applying downstream causally informative approaches, future studies can attempt to isolate specific individual-level mechanisms underpinning rises in depressive symptoms and diagnoses in adolescence.

## Declaration on Use of AI

I have used AI to get some help of R codes and modify some sentences while writing this paper.

## Data Availability

All data produced in the present study are available upon request to Norwegian Institute of Public Health.

## References

1. Thapar A, Collishaw S, Pine DS, Thapar AK. Depression in adolescence. The Lancet. 2012 Mar;379(9820):1056–67. doi:10.1016/S0140-6736(11)60871-4

2. Rohde P, Lewinsohn PM, Klein DN, Seeley JR, Gau JM. Key Characteristics of Major Depressive Disorder Occurring in Childhood, Adolescence, Emerging Adulthood, and Adulthood. Clinical Psychological Science. 2013 Jan;1(1):41–53. doi:10.1177/2167702612457599

3. Weissman MM, Wickramaratne P, Nomura Y, Warner V, Pilowsky D, Verdeli H. Offspring of Depressed Parents: 20 Years Later. AJP. 2006 Jun;163(6):1001–8. doi:10.1176/ajp.2006.163.6.1001

4. Weavers B, Riglin L, Martin J, Anney R, Collishaw S, Heron J, et al. Corrigendum to “Characterising depression trajectories in young people at high familial risk of depression” [J. Affect. Disord. 337 (2023) 66–74 (15 September)]. Journal of Affective Disorders. 2023 Oct;339:1002. doi:10.1016/j.jad.2023.06.039

5. Stephens A, Allardyce J, Weavers B, Lennon J, Jones RB, Powell V, et al. Developing and validating a prediction model of adolescent major depressive disorder in the offspring of depressed parents. Child Psychology Psychiatry. 2023 Mar;64(3):367–75. doi:10.1111/jcpp.13704

6. Kim-Cohen J, Caspi A, Moffitt TE, Harrington H, Milne BJ, Poulton R. Prior Juvenile Diagnoses in Adults With Mental Disorder: Developmental Follow-Back of a Prospective-Longitudinal Cohort. Arch Gen Psychiatry. 2003 Jul 1;60(7):709. doi:10.1001/archpsyc.60.7.709

7. Flint J, Kendler KS. The Genetics of Major Depression. Neuron. 2014 Feb;81(3):484–503. doi:10.1016/j.neuron.2014.01.027

8. Rice F, Harold G, Thapar A. The genetic aetiology of childhood depression: a review. Child Psychology Psychiatry. 2002 Jan;43(1):65–79. doi:10.1111/1469-7610.00004

9. Schoeler T, Duncan L, Cecil CM, Ploubidis GB, Pingault JB. Quasi-experimental evidence on short– and long-term consequences of bullying victimization: A meta-analysis. Psychological Bulletin. 2018 Dec;144(12):1229–46. doi:10.1037/bul0000171

10. Collishaw S. Annual Research Review: Secular trends in child and adolescent mental health. Child Psychology Psychiatry. 2015 Mar;56(3):370–93. doi:10.1111/jcpp.12372

11. Kessler RC, Berglund P, Demler O, Jin R, Merikangas KR, Walters EE. Lifetime Prevalence and Age-of-Onset Distributions of DSM-IV Disorders in the National Comorbidity Survey Replication. Arch Gen Psychiatry. 2005 Jun 1;62(6):593. doi:10.1001/archpsyc.62.6.593

12. Lohoff FW. Overview of the Genetics of Major Depressive Disorder. Curr Psychiatry Rep. 2010 Dec;12(6):539–46. doi:10.1007/s11920-010-0150-6 PubMed PMID: 20848240; PubMed Central PMCID: PMC3077049.

13. Wickramaratne PJ, Greenwald S, Weissman MM. Psychiatric Disorders in the Relatives of Probands With Prepubertal-Onset or Adolescent-Onset Major Depression. Journal of the American Academy of Child & Adolescent Psychiatry. 2000 Nov;39(11):1396–405. doi:10.1097/00004583-200011000-00014

14. Neuman RJ, Geller B, Rice JP, Todd RD. Increased Prevalence and Earlier Onset of Mood Disorders Among Relatives of Prepubertal Versus Adult Probands. Journal of the American Academy of Child & Adolescent Psychiatry. 1997 Apr;36(4):466–73. doi:10.1097/00004583-199704000-00008

15. Puig-Antich J. A Controlled Family History Study of Prepubertal Major Depressive Disorder. Arch Gen Psychiatry. 1989 May 1;46(5):406. doi:10.1001/archpsyc.1989.01810050020005

16. Grimes PZ, Mitchell BL, Thompson KN, Deng Q, Shen X, Wolfe JC, et al. Genome-wide association study of adolescent-onset depression [Internet]. Genetic and Genomic Medicine; 2025 [cited 2025 Dec 10]. Available from: http://medrxiv.org/lookup/doi/10.1101/2025.09.26.25335972 doi:10.1101/2025.09.26.25335972

17. Shorter JR, Pasman JA, Kurvits S, Jangmo A, Naamanka J, Harder A, et al. Genome-wide association analyses identify distinct genetic architectures for early-onset and late-onset depression. Nat Genet. 2025 Nov 13. doi:10.1038/s41588-025-02396-8

18. Lau JYF, Eley TC. Changes in genetic and environmental influences on depressive symptoms across adolescence and young adulthood. Br J Psychiatry. 2006 Nov;189(5):422–7. doi:10.1192/bjp.bp.105.018721

19. Hannigan LJ, Walaker N, Waszczuk MA, McAdams TA, Eley TC. Aetiological Influences on Stability and Change in Emotional and Behavioural Problems across Development: A Systematic Review. Psychopathology Review. 2017 Mar;a4(1):52–108. doi:10.5127/pr.038315

20. Rice F, Riglin L, Thapar AK, Heron J, Anney R, O’Donovan MC, et al. Characterizing Developmental Trajectories and the Role of Neuropsychiatric Genetic Risk Variants in Early-Onset Depression. JAMA Psychiatry. 2019 Mar 1;76(3):306. doi:10.1001/jamapsychiatry.2018.3338

21. Kwong ASF, Morris TT, Pearson RM, Timpson NJ, Rice F, Stergiakouli E, et al. Polygenic risk for depression, anxiety and neuroticism are associated with the severity and rate of change in depressive symptoms across adolescence. Child Psychology Psychiatry. 2021 Dec;62(12):1462–74. doi:10.1111/jcpp.13422

22. Shakeshaft A, Martin J, Dennison CA, Riglin L, Lewis CM, O’Donovan MC, et al. Estimating the impact of transmitted and non-transmitted psychiatric and neurodevelopmental polygenic scores on youth emotional problems. Mol Psychiatry. 2024 Feb;29(2):238–46. doi:10.1038/s41380-023-02319-1

23. Pingault J, Allegrini AG, Odigie T, Frach L, Baldwin JR, Rijsdijk F, et al. Research Review: How to interpret associations between polygenic scores, environmental risks, and phenotypes. Child Psychology Psychiatry. 2022 Oct;63(10):1125–39. doi:10.1111/jcpp.13607

24. Kong A, Thorleifsson G, Frigge ML, Vilhjalmsson BJ, Young AI, Thorgeirsson TE, et al. The nature of nurture: Effects of parental genotypes. Science. 2018 Jan 26;359(6374):424–8. doi:10.1126/science.aan6877

25. Young AS. Estimation of indirect genetic effects and heritability under assortative mating [Internet]. Genetics; 2023 [cited 2025 Nov 10]. Available from: http://biorxiv.org/lookup/doi/10.1101/2023.07.10.548458 doi:10.1101/2023.07.10.548458

26. Askelund AD, Wootton RE, Torvik FA, Lawn RB, Ask H, Corfield EC, et al. Assessing causal links between age at menarche and adolescent mental health: a Mendelian randomisation study. BMC Med. 2024 Apr 12;22(1):155. doi:10.1186/s12916-024-03361-8

27. Bergen SE, Gardner CO, Kendler KS. Age-Related Changes in Heritability of Behavioral Phenotypes Over Adolescence and Young Adulthood: A Meta-Analysis. Twin Res Hum Genet. 2007 Jun 1;10(3):423–33. doi:10.1375/twin.10.3.423

28. Eilertsen EM, Jami ES, McAdams TA, Hannigan LJ, Havdahl AS, Magnus P, et al. Direct and Indirect Effects of Maternal, Paternal, and Offspring Genotypes: Trio-GCTA. Behav Genet. 2021 Mar;51(2):154–61. doi:10.1007/s10519-020-10036-6

29. Cheesman R, Eilertsen EM, Ahmadzadeh YI, Gjerde LC, Hannigan LJ, Havdahl A, et al. How important are parents in the development of child anxiety and depression? A genomic analysis of parent-offspring trios in the Norwegian Mother Father and Child Cohort Study (MoBa). BMC Med. 2020 Dec;18(1):284. doi:10.1186/s12916-020-01760-1

30. Magnus P, Birke C, Vejrup K, Haugan A, Alsaker E, Daltveit AK, et al. Cohort Profile Update: The Norwegian Mother and Child Cohort Study (MoBa). Int J Epidemiol. 2016 Apr;45(2):382–8. doi:10.1093/ije/dyw029

31. Brandlistuen RE, Kristjansson D, Alsaker E, Valen R, Birkeland E, Røyrvik EC, et al. Cohort Profile Update: The Norwegian Mother, Father and Child Cohort (MoBa). International Journal of Epidemiology. 2025 Aug 18;54(5):dyaf139. doi:10.1093/ije/dyaf139

32. Paltiel L, Anita H, Skjerden T, Harbak K, Bækken S, Nina Kristin S, et al. The biobank of the Norwegian Mother and Child Cohort Study – present status. Nor J Epidemiol. 2014 Dec 22;24(1–2). doi:10.5324/nje.v24i1-2.1755

33. Corfield EC, Shadrin AA, Frei O, Rahman Z, Lin A, Athanasiu L, et al. The Norwegian Mother, Father, and Child cohort study (MoBa) genotyping data resource: MoBaPsychGen pipeline v.1 [Internet]. 2022 [cited 2025 Sep 11]. Available from: http://biorxiv.org/lookup/doi/10.1101/2022.06.23.496289 doi:10.1101/2022.06.23.496289

34. Angold, A., Costello, E. J., & Messer, S. C. (1996). Development of a short questionnaire for use in epidemiological studies of depression in children and adolescents. Vol. 5. 1996 Dec 1;5(4):237–49.

35. McIntosh AM, Lewis CM, Mark J Adams for the Psychiatric Genomics Consortium Major Depressive Disorder Working Group. Genome-wide study of half a million individuals with major depression identifies 697 independent associations, infers causal neuronal subtypes and biological targets for novel pharmacotherapies [Internet]. 2024 [cited 2024 Nov 13]. Available from: http://medrxiv.org/lookup/doi/10.1101/2024.04.29.24306535 doi:10.1101/2024.04.29.24306535

36. Hughes AM, Torvik FA, Van Bergen E, Hannigan LJ, Corfield EC, Andreassen OA, et al. Parental education and children’s depression, anxiety, and ADHD traits, a within-family study in MoBa. npj Sci Learn. 2024 Jul 18;9(1):46. doi:10.1038/s41539-024-00260-8

37. Riglin L, Thapar AK, Leppert B, Martin J, Richards A, Anney R, et al. Using Genetics to Examine a General Liability to Childhood Psychopathology. Behav Genet. 2020 Jul;50(4):213–20. doi:10.1007/s10519-019-09985-4 PubMed PMID: 31828458; PubMed Central PMCID: PMC7355267.

38. Askelund AD, Hegemann L, Allegrini AG, Corfield EC, Ask H, Davies NM, et al. The Genetic Architecture of Differentiating Behavioral and Emotional Problems in Early Life. Biological Psychiatry. 2025 Jun;97(12):1163–74. doi:10.1016/j.biopsych.2024.12.021

39. Demontis D, Walters GB, Athanasiadis G, Walters R, Therrien K, Nielsen TT, et al. Genome-wide analyses of ADHD identify 27 risk loci, refine the genetic architecture and implicate several cognitive domains. Nat Genet. 2023 Feb;55(2):198–208. doi:10.1038/s41588-022-01285-8

40. Strom NI, Verhulst B, Bacanu SA, Cheesman R, Purves KL, Gedik H, et al. Genome-wide association study of major anxiety disorders in 122,341 European-ancestry cases identifies 58 loci and highlights GABAergic signaling [Internet]. Genetic and Genomic Medicine; 2024 [cited 2025 Nov 10]. Available from: http://medrxiv.org/lookup/doi/10.1101/2024.07.03.24309466 doi:10.1101/2024.07.03.24309466

41. Privé F, Arbel J, Vilhjálmsson BJ. LDpred2: better, faster, stronger. Schwartz R, editor. Bioinformatics. 2021 Apr 1;36(22–23):5424–31. doi:10.1093/bioinformatics/btaa1029

42. Buuren SV, Groothuis-Oudshoorn K. mice: Multivariate Imputation by Chained Equations in R. J Stat Soft. 2011;45(3). doi:10.18637/jss.v045.i03

43. R Core Team. R: A language and environment for statistical computing [Internet]. Vienna, Austria: R Foundation for Statistical Computing; 2024. Available from: https://www.R-project.org/

44. Benjamini Y, Hochberg Y. Controlling the False Discovery Rate: A Practical and Powerful Approach to Multiple Testing. Journal of the Royal Statistical Society Series B: Statistical Methodology. 1995 Jan 1;57(1):289–300. doi:10.1111/j.2517-6161.1995.tb02031.x

45. Hannigan LJ, Corfield EC, Askelund AD, Askeland RB, Hegemann L, Jensen P, et al. phenotools: an R package to facilitate efficient and reproducible use of phenotypic data from MoBa and linked registry sources in the TSD environment [Internet]. 2023 Sep 6 [cited 2023 Nov 6]. Available from: 10.17605/OSF.IO/6G8BJ

46. Mostafavi H, Harpak A, Agarwal I, Conley D, Pritchard JK, Przeworski M. Variable prediction accuracy of polygenic scores within an ancestry group. eLife. 2020 Jan 30;9:e48376. doi:10.7554/eLife.48376

47. Rice F. The genetics of depression in childhood and adolescence. Curr Psychiatry Rep. 2009 Apr;11(2):167–73. doi:10.1007/s11920-009-0026-9

48. Veller C, Coop GM. Interpreting population– and family-based genome-wide association studies in the presence of confounding. Moorjani P, editor. PLoS Biol. 2024 Apr 11;22(4):e3002511. doi:10.1371/journal.pbio.3002511

49. Howe LJ, Nivard MG, Morris TT, Hansen AF, Rasheed H, Cho Y, et al. Within-sibship genome-wide association analyses decrease bias in estimates of direct genetic effects. Nat Genet. 2022 May;54(5):581–92. doi:10.1038/s41588-022-01062-7

50. 23andMe Research Team, COGENT (Cognitive Genomics Consortium), Social Science Genetic Association Consortium, Lee JJ, Wedow R, Okbay A, et al. Gene discovery and polygenic prediction from a genome-wide association study of educational attainment in 1.1 million individuals. Nat Genet. 2018 Aug;50(8):1112–21. doi:10.1038/s41588-018-0147-3

51. Biele G, Gustavson K, Czajkowski NO, Nilsen RM, Reichborn-Kjennerud T, Magnus PM, et al. Bias from self selection and loss to follow-up in prospective cohort studies. Eur J Epidemiol. 2019 Oct;34(10):927–38. doi:10.1007/s10654-019-00550-1

52. Bilghese M, Manansala R, Jaishankar D, Jala J, Benjamin DJ, Kimball M, et al. A General Approach to Adjusting Genetic Studies for Assortative Mating. bioRxiv. 2023 Sep 5;2023.09.01.555983. doi:10.1101/2023.09.01.555983 PubMed PMID: 37732240; PubMed Central PMCID: PMC10508715.

53. Liskola K, Raaska H, Lapinleimu H, Lipsanen J, Sinkkonen J, Elovainio M. The effects of maternal depression on their perception of emotional and behavioral problems of their internationally adopted children. Child Adolesc Psychiatry Ment Health. 2021 Dec;15(1):41. doi:10.1186/s13034-021-00396-0

54. Van Der Toorn SLM, Huizink AC, Utens EMWJ, Verhulst FC, Ormel J, Ferdinand RF. Maternal depressive symptoms, and not anxiety symptoms, are associated with positive mother–child reporting discrepancies of internalizing problems in children: a report on the TRAILS Study. Eur Child Adolesc Psychiatry. 2010 Apr;19(4):379–88. doi:10.1007/s00787-009-0062-3

55. Zhang X, Grove J, Gu Y, Buus CK, Nielsen LK, Neufeld SAS, et al. Polygenic and developmental profiles of autism differ by age at diagnosis. Nature. 2025 Oct 30;646(8087):1146–55. doi:10.1038/s41586-025-09542-6

