## Supplementary for "Estimating direct and indirect genetic effects on variation in depressive symptoms in early adolescence: a trio PGS analysis in the MoBa cohort": DIGE Supplementary material_MedRxiv.pdf

### Supplementary Methods 1

**Outcome: Short Mood and Feelings Questionnaire (SMFQ): Description of original scale**

The Mood and Feelings Questionnaire (MFQ; Angold & Costello, 1987) is a 32-item questionnaire based on DSM-III-R criteria for depression. The MFQ consists of a series of descriptive phrases regarding how the subject has been feeling or acting recently. A 13-item short form was developed, based on the discriminating ability between the depressed and nondepressed (1). Both parent and child-report forms are available. The parent version is used in the MoBa 8-year questionnaire. The short mood and feeling questionnaire (SMFQ) assess depressive symptoms experienced over the past two weeks, using a three-point Likert scale (Not True, Sometimes, True) for each item. In this study mothers responded to 13 items at age 8 and 6 items at age 14 while children reported 13 items only at age 14. It is a condensed version of the full Mood and Feelings Questionnaire (MFQ), designed to be a rapid and valid screening tool for depression in various settings.

**Exposures: Polygenic Score (PGS) overview**

PGS are individual-level measures of genetic liability computed as a weighted sum of the effect alleles across single-nucleotide polymorphisms (SNPs) in a person's genome. The weights are derived based on the strength of SNP-trait associations established in a prior Genome Wide Association Studies (GWAS) for a given trait or disorder (2,3). We constructed PGS using summary statistics from the recent Psychiatric Genomics Consortium GWAS of major depressive disorder. Next, to account for the scenario that this PGS, which is based on GWAS in adults, does not capture all of the genetic liability (4) that is relevant to child and adolescent depressive symptoms, and because of the genetic overlap across traits, we additionally included PGS for 3 other traits that have been demonstrated to explain variation in childhood emotional problems namely: ADHD. Anxiety and EA.

- Major depressive disorder (MDD) PGS calculated based on the latest Psychiatric Genomics Consortium (PGC) GWAS (5), for which summary statistics excluding the Norwegian mother, father and child cohort study (MoBa) have been supplied to us directly.
- ADHD PGS calculated based on summary statistics from the GWAS conducted by Demontis et al., 2023 (6).
- Anxiety PGS calculated based on the most updated PGC GWAS, for which summary statistics excluding MoBa have been supplied to us directly (7).
- Educational attainment PGS calculated using the GWAS conducted by Okbay A, et al (8), for which summary statistics excluding MoBa have been supplied to us directly.

We computed PGS using LDpred2 (9), with the option 'LDPred2-auto' . The recommended quality control steps (10) were followed using an established pipeline (9) that includes restricting variants to an extended set of HAPMAP3+ variants (1.4 million SNPs). Additionally, only well-imputed SNPs in MoBa (INFO score >0.95) were included. Precomputed LD matrices from UK Biobank were used as the reference LD panel (11). PGSs were regressed on the first 20 genomic principal components (PCs), genotype and imputation batch then standardized to have a mean of 0 and a standard deviation of 1 prior to inclusion in analyses.

Table S1- Preregistration Deviation table

<table>
<tr><th colspan="6">Deviations</th></tr>
<tr><th>#</th><th colspan="2">Details</th><th>Original Wording</th><th>Deviation Description</th><th>Reader Impact</th></tr>
<tr><td rowspan="3">1</td><td>Type</td><td>Analysis</td><td rowspan="3">In the preregistration it was stated sensitivity analysis by fitting a model using “13 mother-reported Short mood and feeling items at age 8, 13 child-reported items at age 14”</td><td rowspan="3">The model using the 13 child-reported items failed to converge. Therefore, we were unable to complete this planned sensitivity analysis.</td><td rowspan="3">This deviation has minimal impact on the interpretation of the main findings. The sensitivity analysis was intended to provide additional insight into potential reporter bias, but its absence does not affect the primary results. The deviation is solely due to model non-convergence.</td></tr>
<tr><td>Reason</td><td>Plan not possible</td></tr>
<tr><td>Timing</td><td>After data access</td></tr>
</table>

<table>
<tr><th colspan="6">Unregistered Steps</th></tr>
<tr><th>#</th><th colspan="2">Details</th><th>Original Wording</th><th>Unregistered Step Description</th><th>Reader Impact</th></tr>
<tr><td rowspan="2">1</td><td>Type</td><td>Analysis</td><td rowspan="2">Our primary analyses use only the 6 depressive symptom items which are consistent across time and reporter to avoid confounding developmental change with measurement changes (i.e., different</td><td rowspan="2">In the description, we did not specify whether the sensitivity analyses would be conducted as independent models at ages 8 and 14, or as a model of emerging depressive symptoms at age 14 relative to age 8.</td><td rowspan="2">This imprecisely registered part of the sensitivity analysis plan has negligible impact on readers’ interpretation of the main findings.</td></tr>
<tr><td>Timing</td><td>After data access</td></tr>
</table>

| | | | |
|---|---|---|---|
| | | items or different reporters across measurement occasions). However, with this confounding as a caveat, we also conduct the following sensitivity analyses:<br>- Sensitivity analysis 1: All mother reported items (13 items at age 8, 6 items at age 14)<br>- Sensitivity analysis 2: 13 mother-reported items at age 8, 13 child-reported items at age 14 | Our intention was to run **independent models at ages 8 and 14** rather than modelling emerging symptoms from 8 to 14. This decision is based on the fact that effects from a longitudinal model with either alternative outcome (i.e., using 7 additional mother-reported items at age 8 or using child-instead of -mother reported depressive symptoms at age 14 ) would not be reasonably interpretable as effects on emerging depressive symptoms in adolescence, as developmental change (i.e., wave of measurement) would be confounded with changes to the measure of the outcome. |

**Explanation of Choosing Covariate**

In gene environment studies it is known that only passive gene environment correlation ( rGE) is confounding while active and evocative rGE are part of the causal pathway from genotype to phenotype.

**Passive rGE**

**e.g.**

- Parents pass on genes *and* environments.

- Example: Educated parents pass both:
    - genes related to cognitive ability
    - a cognitively stimulating home

This creates confounding, because environment and genotype are correlated *without* the individual doing anything.

**Passive rGE** : Genotype and environment are correlated because of shared causes (e.g., parents)

**Active & evocative rGE** (causal pathway)

- **Active rGE**: a child seeks environments based on their genotype
- **Evocative rGE**: Others respond to their genotype

These are not confounders — they are mechanisms through which genes affect outcomes.

**Active & evocative rGE:** Genotype and environment are correlated because the focal person *selects, creates, or evokes* their environment based on their genotype (e.g., peers, partners, neighborhoods). Crucially, active and evocative gene-environment correlations are *not confounding*. They are part of the *causal pathway from genotype to phenotype* and are therefore mediators of the genetic effect.

In a trio PGS model parental PGS, parental educational attainment and household income acts as a control for passive rGE confounding which *could affect* the true association of the *child trait and their genotype* (direct genetic effect).

Mediators should *NOT* be controlled if we want the total genetic effect. If we want to control for mediators we may commit overadjustment, we are also blocking part of the causal pathway and underestimating the true genetic effect.

Table S2: List of covariates

| Variable | Data type | Measure | Source | Notes |
|---|---|---|---|---|
| Child sex | Binary | M/F | MBRN | |
| Parity | Count | In number of previous births | MBRN | |
| Birth year | Continuous (standardized) | In years | MBRN | |
| Mother's age at childbirth | Continuous | In years | MBRN | |
| Father's age at childbirth | Continuous | In years | MBRN | |
| Household income at child age 8 | Ordinal | Composite measure | Statistics Norway (SSB) | Maternal income + paternal income |
| **Educational attainment** | | | | |
| Father's educational attainment at age 8 | Ordinal | Highest completed education in number of years | SSB | |
| Mother's educational attainment at age 8 | Ordinal | Highest completed education in number of years | SSB | |

*Medical Birth Registry of Norway (MBRN)

Table S3: Available data for each variable from the total genotyped 42,059 sample.

| Variables | N(%) | Mean | Std.Dev. | Min | Max | Missing n(%) |
|---|---|---|---|---|---|---|
| **Imputed variables** | | | | | | |
| Depressive symptom score at 8 on CCA | 18,572 (44.05) | 6.98 | 1.29 | 0 | 18 | 23,532 (55.95) |
| Dep symp score at 14 on CCA | 11588 (27.55) | 7 | 1.85 | 6 | 18 | 30, 471 (72.45) |
| HH income | 41298 (98.19) | 856617.8 | 325386.5 | <250000 | >3000000 | 761 (1.81) |
| Mothers edu | 41,969 (99.79) | 16 (median) | - | 0 | 21 | 90 ((0.214)) |
| Fathers edu | 41,931 (99.70) | 16 (median) | - | 0 | 21 | 128(0.304) |
| **Variables in the analytic model** | | | | | | |
| MDD PGS child | 42,059 | -0.0222 | 0.9979 | -3.785 | 4.005 | 0 |
| MDD PGS mothers | 42,059 | -0.0076 | 0.997 | -4.110 | 4.287 | 0 |
| MDD PGS fathers | 42,059 | -0.0086 | 1.001 | -3.881 | 3.940 | 0 |
| ADHD PGS child | 42,059 | -0.0304 | 1.0005 | -4.1465 | 4.594 | 0 |
| ADHD PGS mother | 42,059 | -0.006 | 1.003 | -3.9023 | 4.345 | 0 |
| ADHD PGS father | 42,059 | -0.002788 | 0.998289 | -4.547596 | 3.992079 | 0 |
| Anxiety PGS child | 42,059 | -0.0123 | 0.9979 | -4.2908 | 4.1481 | 0 |
| Anxiety PGS mother | 42,059 | -0.00377 | 0.989778 | -4.2142 | 4.31123 | 0 |
| Anxiety PGS father | 42,059 | -0.00254 | 1.002455 | -4.1675 | 4.478742 | 0 |
| EA PGS child | 42,059 | 0.01933 | 1.01034 | -4.01528 | 4.25980 | 0 |
| EA PGS mother | 42,059 | 0.03023 | 0.99137 | -3.93543 | 4.17172 | 0 |
| EA PGS father | 42,059 | 0.009069 | 1.001639 | -3.9075 | 4.286526 | 0 |
| Parity | 42,059 | 1 (median) | - | 0 | 4 | 0 |
| Mothers age at birth | 42,059 | 30.16 | 4.4 | 16 | 46 | 0 |
| Fathers age at birth | 42,055 | 32.56 | 5.14 | 18 | 60 | 4 |
| Child sex | | | | | | |
| Male | 21451 (51.00) | - | - | - | - | 0 |
| Female | 20608 (49.00) | - | - | - | - | 0 |

*Note: PGS=polygenic score; MDD=major depressive disorder, ADHD=Attention-deficit/hyperactivity disorder; EA=educational attainment.*

Table S4: Comparison of the full sample (before restricted to genotyped n=113,875) and analytic sample n=42,059.

| **Full sample (N=113875)** | | | | | **Analytic sample (42,059)** | | | |
|---|---|---|---|---|---|---|---|---|
| **Variables** | **N(%)** | **Mean (S.D)** | **Missing n(%)** | **Min (Max)** | **N(%)** | **Mean (S.D)** | **Missing n(%)** | **Min (Max)** |
| **Depressive symptoms score** | | | | | | | | |
| At age 8 | 42,625 (37.43) | 6.99 (1.32) | 71250 (62.56) | | 18,572 (44.05) | 6.98 (1.29) | 23,532 (55.95) | |
| At age 14 | 25,778 (22.64) | 7.1 (1.88) | 88097 (77.36) | | 11588 (27.55) | 7.0 (1.85) | 30, 471 (72.45) | |
| **Sex** | | | | | | | | |
| Male | 58009 (51.15) | - | 476 | - | 21451 (51.00) | - | 0 | |
| Female | 55176 (48.65) | - | | - | 20608 (49.00) | - | 0 | |
| Not specified | 174 (0.15) | - | | - | - | - | - | |
| Uncertain | 40 (0.04) | - | | - | - | - | - | |
| **Covariates** | | | | | | | | |
| HH income | 110,557 (97.09) | 828197.6 (339789.5) | 3318 (2.91) | 170,001 (3460001) | 41298 (98.19) | 856617.8 (325386.5) | 761 (1.81) | <250000 (>3000000) |
| Mother edu | 112,974 | 16 (median) | 901 | 0 (21) | 41,969 (99.79) | 16 (median) | 90 (0.214) | 0 (21) |
| Father edu | 86,969 | 13 (median) | 26906 | 0 (21) | 41,931 (99.70) | 16 (median) | 128 (0.304) | 0 (21) |
| Mothers age at birth | | 30.15 (4.6) | 476() | | 42,059 | 30.16 (4.4) | | 16 (46) |
| Fathers age at birth | | 32.74 (5.47) | 994() | | 42,055 | 32.56 (5.14) | 4 | 18 (60) |

Table S5. Mother reported items at age 8 and 14.

| Mother reported items at age 8 | | | | | | | |
|---|---|---|---|---|---|---|---|
| **Items** | **Response** | **n** | **%** | **Items** | **Response** | **n** | **%** |
| Felt miserable or unhappy | Not true | 10,491 | 24.9 | Thought s/he could never be as good as other kids | Not true | 16,535 | 39.3 |
| | Sometimes true | 7,908 | 18.8 | | Sometimes true | 2,118 | 5.0 |
| | True | 281 | 0.7 | | True | 80 | 0.2 |
| | NA | 23,379 | 55.6 | | NA | 23,326 | 55.5 |
| Felt so tired that s/he just sat around and did nothing | Not true | 16,859 | 40.1 | Felt lonely | Not true | 16,820 | 40.0 |
| | Sometimes true | 1,788 | 4.3 | | Sometimes true | 1,854 | 4.4 |
| | True | 99 | 0.2 | | True | 57 | 0.1 |
| | NA | 23,313 | 55.4 | | NA | 23,328 | 55.5 |
| Was very restless | Not true | 14,760 | 35.1 | Thought nobody really loved him/her | Not true | 17,722 | 42.1 |
| | Sometimes true | 3,632 | 8.6 | | Sometimes true | 981 | 2.3 |
| | True | 340 | 0.8 | | True | 32 | 0.1 |
| | NA | 23,327 | 55.5 | | NA | 23,324 | 55.5 |
| Didn’t enjoy anything | Not true | 16,942 | 40.3 | Felt s/he was a bad person | Not true | 17,710 | 42.1 |
| | Sometimes true | 1,685 | 4.0 | | Sometimes true | 999 | 2.4 |
| | True | 108 | 0.3 | | True | 22 | 0.1 |
| | NA | 23,324 | 55.4 | | NA | 23,328 | 55.5 |
| Felt s/he was no good anymore | Not true | 17,023 | 40.5 | Felt s/he did everything wrong | Not true | 15,933 | 37.9 |
| | Sometimes true | 1,659 | 3.9 | | Sometimes true | 2,732 | 6.5 |
| | True | 51 | 0.1 | | True | 56 | 0.1 |
| | NA | 23,326 | 55.5 | | NA | 23,338 | 55.5 |
| Cried a lot | Not true | 17,138 | 40.7 | Found it hard to think/concentrate | Not true | 15,201 | 36.1 |
| | Sometimes true | 1,562 | 3.7 | | Sometimes true | 3,216 | 7.6 |
| | True | 51 | 0.1 | | True | 291 | 0.7 |
| | NA | 23,308 | 55.4 | | NA | 23,351 | 55.5 |
| Hated himself/herself | Not true | 17,810 | 42.3 | | | | |
| | Sometimes true | 908 | 2.2 | | | | |
| | True | 26 | 0.1 | | | | |
| | NA | 23,315 | 55.4 | | | | |

| Mother reported items at age 14 | | | | | | | |
|---|---|---|---|---|---|---|---|
| **Items** | **Response** | **n** | **%** | **Items** | **Response** | **n** | **%** |
| Was sad/unhappy (UM 178) | Not true | 8,912 | 21.2 | Thought he/she could never be as good as other children (181) | Not true | 10,137 | 24.1 |
| | Sometimes true | 2,576 | 6.1 | | Sometimes true | 1,382 | 3.3 |
| | True | 320 | 0.8 | | True | 249 | 0.6 |
| | NA | 30,251 | 71.9 | | NA | 30,291 | 72.0 |
| Felt so tired that he/she just sat there doing nothing (UM 179) | Not true | 8,682 | 20.6 | Thought no one really liked him/her (182) | Not true | 11,065 | 26.3 |
| | Sometimes true | 2,738 | 6.5 | | Sometimes true | 611 | 1.5 |
| | True | 397 | 0.9 | | True | 84 | 0.2 |
| | NA | 30,242 | 71.9 | | NA | 30,299 | 72.0 |
| Felt worthless (180) | Not true | 10,163 | 24.2 | Thought he/she did everything wrong (183) | Not true | 10,278 | 24.4 |
| | Sometimes true | 1,371 | 3.3 | | Sometimes true | 1,341 | 3.2 |
| | True | 247 | 0.6 | | True | 136 | 0.3 |
| | NA | 30,278 | 72.0 | | NA | 30,304 | 72.1 |

### Supplementary Methods 2

**Missing mechanism test**

Mother reported items at age 8 and 14 and child reported at age 14 were tested for missing mechanisms using Little's MCAR test. This test checks whether the pattern of missingness is random (12). It's a multivariate test based on comparing means and covariances between observed and missing groups. The null hypothesis provides evidence that variables align with missing completely at random (MCAR) mechanism. The test indicated that mother reported items at both age with p-values <0.001 and child reported items with P-value 0.00285 evidenced not MCAR.

In addition, missing data indicators were created for each item, and a binary logistic regression was fitted to assess whether the missingness has association with variables in the model called missing at random (MAR) (13). All mother reported items at age 8 are associated with child ADHD PGS, Mother EA PGS, father anxiety PGS, mother MDD PGS, both parent educational attainment, parity, childbirth year, both parental age at childbirth, except one of the items which didn't show association with Parity. While mother reported items at age 14 were associated with Mother MDD PGS, Parity, child sex, both parent educational attainment, birth year, HH income and fathers age at childbirth. Therefore, we can conclude that the mother reported items missingness at age 8 and 14 were associated with observed data which is evidence of Missing At Random (MAR). Similarly, all child reported items at age 14 missingness were consistently associated with child ADHD PGS, child EA PGS, birth year, HH income, both parent educational attainment, child sex, mother and child MDD PGS and fathers age at birth. Therefore, we can assume the possibility of MAR.

Besides, a comparison of mean polygenic scores for each exposure PGS between complete and incomplete depressive symptoms was performed (*Table S6)*. The t-test indicated that, there is significant mean difference on maternal MDD, ADHD, Anxiety and EA PGS between missing and observed participants with P-value <0.001 for all PGS.. Differences in average PGS between missing and observed groups indicate that genetic liability may be related to missingness patterns. This supports the use of imputation methods that assume data are missing at random (MAR).

Table S6: Comparing mean of PGS between observed and unobserved based on depressive symptoms

| Role | PGS | Est. | Stat. | p-value | Mean of missing | S.d of missing | Mean of observed | S.d of observed |
|---|---|---|---|---|---|---|---|---|
| Child | MDD | -0.093 | -9.536 | <0.001 | 0.021 | 0.998 | -0.073 | 0.995 |
| | ADHD | -0.097 | -9.884 | <0.001 | 0.029 | 0.999 | -0.068 | 1.000 |
| | Anxiety | -0.042 | -4.238 | <0.001 | 0.006 | 0.996 | -0.036 | 1.000 |
| | EA | 0.143 | 14.416 | <0.001 | -0.043 | 1.011 | 0.100 | 1.004 |
| Mother | MDD | -0.129 | -13.230 | <0.001 | 0.049 | 0.996 | -0.081 | 0.994 |
| | ADHD | -0.093 | -9.435 | <0.001 | 0.034 | 1.002 | -0.059 | 1.002 |
| | Anxiety | -0.058 | -6.013 | <0.001 | 0.022 | 0.994 | -0.037 | 0.984 |
| | EA | 0.163 | 16.843 | <0.001 | -0.041 | 0.994 | 0.122 | 0.980 |

#### Selective attrition

Table S7: Selective attrition: 8-yr SMFQ scores by availability of 14-year data

| **data_14yr** | **N** | **mean_8yr_smfq** | **SE** |
|---|---|---|---|
| Dropped out | 9707 | 7.032 | 0.014 |
| Stayed in | 8820 | 6.917 | 0.013 |

*Note: Selective attrition based on depressive symptom scores can only be evaluated among participants with data at age 8. Consequently, there is a discrepancy of 2,768 participants between the total in this table and the number reported at age 14 in Table S4, attributable to individuals who were not assessed at age 8 but contributed data at age 14.*

Table S8: Selective attrition: polygenic scores

| Family member | PGS trait | Baseline only - mean | Baseline only - SE | 8yr only - mean | 8yr only - SE | Complete - mean | Complete - SE | Mean diff (complete-baseline) |
|---|---|---|---|---|---|---|---|---|
| Child | Anxiety | 0.010 | 0.007 | -0.019 | 0.01 | -0.055 | 0.011 | |
| | Educational attainment | -0.061 | 0.007 | 0.042 | 0.01 | 0.164 | 0.011 | |
| | MDD | 0.028 | 0.007 | -0.038 | 0.01 | -0.113 | 0.011 | |
| | ADHD | 0.038 | 0.007 | -0.029 | 0.01 | -0.110 | 0.011 | |
| Mother | Anxiety | 0.025 | 0.007 | -0.021 | 0.01 | -0.053 | 0.010 | |
| | Educational attainment | -0.058 | 0.007 | 0.076 | 0.01 | 0.174 | 0.010 | 0.232 |
| | MDD | 0.059 | 0.007 | -0.054 | 0.01 | -0.111 | 0.010 | -0.17 |
| | ADHD | 0.044 | 0.007 | -0.022 | 0.01 | -0.099 | 0.011 | |
| Father | Anxiety | 0.002 | 0.007 | 0.004 | 0.01 | -0.014 | 0.011 | |
| Father | Educational attainment | -0.044 | 0.007 | 0.030 | 0.01 | 0.095 | 0.011 | |
| Father | MDD | 0.009 | 0.007 | -0.016 | 0.01 | -0.048 | 0.011 | |
| Father | ADHD | 0.026 | 0.007 | -0.017 | 0.01 | -0.054 | 0.011 | |

*Complete=those participants stayed at both age 8 and 14. ; Baseline – participants enrolled in the study but who didn't even stay at child age 8.*

Genetic liabilities to anxiety, major depressive disorder (MDD), and ADHD decreased among participants who remained in the study over time, as indicated by a shift from positive mean polygenic score (PGS) values at baseline to negative values at ages 8 and 14. In contrast, genetic liability to educational attainment increased over time (Table S8). These patterns suggest that participants who remained in the study had lower genetic risk for psychiatric traits and relatively higher genetic propensity for educational attainment compared with those who dropped out.

### Supplementary Methods 3

#### **Multilevel multiple imputation**

##### Assigning predictors

All variables (including family members PGS) that appear in the analysis model were included as predictors in multiple imputation models in line with the recommendation to impute based on all variables used in the analytic model (14,15). To reduce computational complexity, no auxiliary variables were included.

All imputed variables and their potential selected predictors correlation were checked and the level of correlation (medium to strong) indicate good predictors (16) . In this study, those variables with **(r>0.2)** with the incomplete variable were used as a predictor for the respective incomplete variables. Weakly associated variables with the incomplete variable were removed from being a predictor. Exceptionally, the weak correlation observed for child and mother reported items were kept to predicting each other regardless of their correlation intensity.

**Specification of MMI models** using MICE

MMI models were used because of the inherent structure of the data, with repeated measures (repeated measures of depressive symptoms at age 8 and 14) nested within individuals, and individuals nested within families (hildren nested within family).

The levels were organized as follows:

1. **Level 1** (Repeated Measures): representing repeated observations of depressive symptoms over time for each individual. At this level—mother reported SMFQ items at age 8 and 14 are included. Hence, we used 2l.pmm method from MICE.
2. **Level 2** (Individual/Child Level): This level represents time-invariant variables (e.g., gender of the child, birth order) and variables related to the child that are measured once could be included. For this study —child reported SMFQ items at age 14 (the only child reported measure in our study), child related variables such sex, child PGS were classified at level 2. We tried to apply 2lonly.pmm method from mice but the imputation couldn't converge. Hence, we finally applied pmm method to impute level 2 missing variables.
3. **Level 3** (Family Level): This level captures mothers and fathers related variables, such as genetic and environmental factors, socioeconomic status, parent PGS. This level variables that had missingness were parent education and household income. Hence, we applied 2lonly.pmm method from MICE.

This hierarchical structure allows the algorithm to take the information of each level as per the assignment of the method and predictors.

Since household income and parental education variables were taken at age 8 of the child. Therefore, for families with more than one child we took the average household income and the highest parental education before imputation.

We have followed the recommendation of model specification to avoid biased estimate. In the interaction terms in the model for e.g (age*PGS), it is known that if none of the interaction terms (for e.g age and PGS) *has missingness, then no need to include such interaction terms in the imputation model.* The interaction should be included if there is missingness in one or both of interaction terms (17). Hence, for this study, all the interaction terms are fully observed for which we haven't included interaction in the imputation model.

**Number of imputations**

Mitigating Monte Carlo error from small samples.

In small samples Monte Carlo variation in imputations can influence results.

- MI involves **simulation**: drawing random values from predictive distributions to replace missing data (18).
- In small samples, random variation in those draws (Monte Carlo error) can **add instability** to the results. That doesn't necessarily mean bias, but it means estimates may **jump around more** between imputed datasets, or if you rerun MI. (19)
- This is why it is emphasized running enough imputations often $m$ = 20–100 or similar amount as the missing proportion in the data so that Monte Carlo error is small compared to sampling variability (20).

*Therefore, in this study the number of imputations used was 75, which is similar to the amount of missingness proportion at the item level.*

**Quality of imputed data**

The quality of the imputed data was checked using density plots (Supplementary Fig S1, S2 and S3) which indicates that the distribution of the observed and imputed items (mother reported SMFQ 6 items at age 8 and 14, and child reported items at age 14) was comparable meaning the imputation process preserved the underlying data structure (19). *We can see that the imputation quality aligns with the recommendation.*

Figure S1. The distribution of observed and imputed data at age 8-mother reported 6 items.

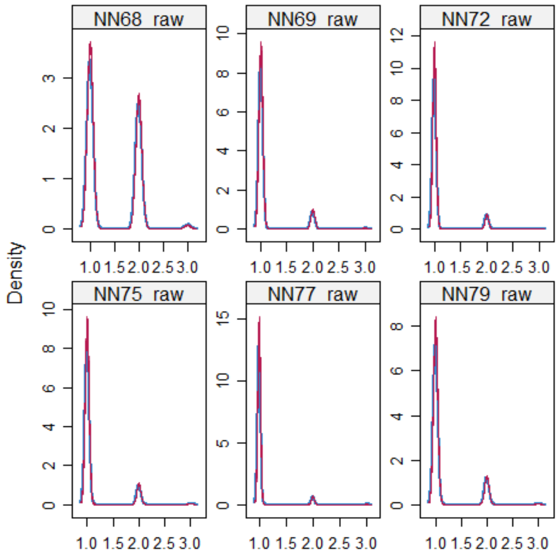

Figure S2. The distribution of the observed and imputed data at age 14-mother reported 6 items.

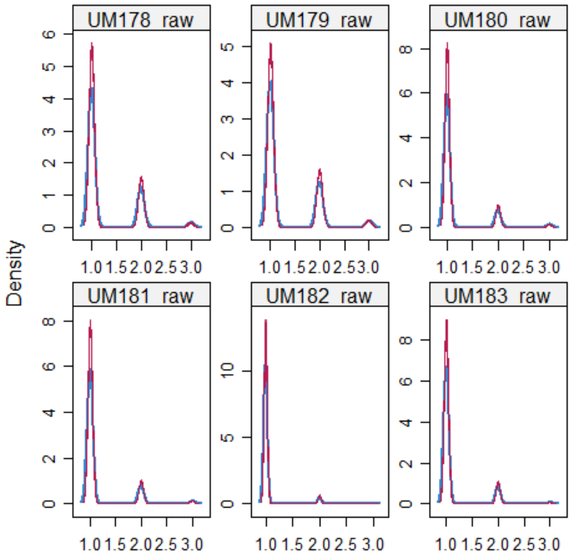

Figure S3: The distribution of observed and imputed data at age 14-mother reported 6 items.

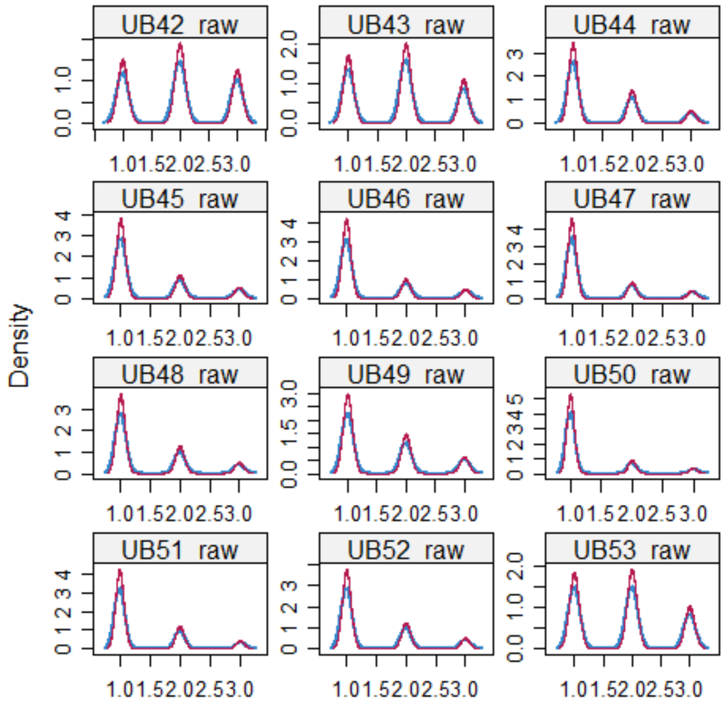

#### Supplementary Methods 4

##### *Analytic Models*

We aimed to estimate the associations of children's genetic liabilities – as well as those of their parents – and the variation of depressive symptoms in early adolescence. For each genetic liability (MDD, anxiety, ADHD, EA) measured by PGS, we constructed a generalized linear mixed model, predicting depressive symptoms while accounting for within-individual level (across time) and within-family level (across siblings) variation. By incorporating main effects of PGS for each member of the mother-father-child trio, as well as their interaction term with wave of measurement, we were able to estimate the direct and indirect genetic effects on both stable depressive symptoms across waves, and adolescent-specific depressive symptoms (variation at age 14 compared to age 8).

Step I: estimating direct and indirect genetic effects at age 8

For each PGS (**MDD, anxiety, ADHD, EA**), we conducted bivariate and a trio approach with generalized linear mixed models, to account for family level variation (siblings), to estimate the direct and indirect genetic effects on depressive symptoms at age 8.

a. *Bivariate model to estimate genetic effects on depressive symptom at age 8*

depression_age8 ~ child_PGS + (1|family_ID)
depression_age8 ~ mother_PGS+ (1|family_ID)
depression_age8 ~ father_PGS+ (1|family_ID)

b. *Trio model to estimate genetic effects on depressive symptom at age 8*

depression_age8 ~ child_PGS+ mother_PGS + father_PGS + (1|family_ID)

c. *Adjusting for covariates*

depression_age8 ~ child_PGS + mother_PGS + father_PGS + parity + sex + HHincome + mother_edu + father_edu + birth_year + maternal_age + paternal_age + (1|family_ID)

Step II: estimating direct and indirect genetic effects at age 14

For each PGS (**MDD, anxiety, ADHD, EA**), we conducted bivariate and a trio approach with generalized linear mixed models, to account for family level variation (siblings), to estimate the direct and indirect genetic effects on depressive symptoms at age 14.

a. *Bivariate model to estimate genetic effects on depressive symptom at age 14*

depression_age14 ~ child_PGS + (1|family_ID)
depression_age14 ~ mother_PGS+ (1|family_ID)
depression_age14 ~ father_PGS+ (1|family_ID)

b. *Trio model to estimate genetic effects on depression at age 14*

depression_age14 ~ child_PGS + mother_PGS + father_PGS+ (1|family_ID)

c. *Adjusting for covariates*

depression_age14 ~ child_PGS + mother_PGS + father_PGS + parity + sex + HHincome + mother_edu + father_edu + birth_year + maternal_age + paternal_age + (1|family_ID)

**<u>Step III: estimating direct and indirect genetic effects on emerging variation in depressive symptoms in early adolescence</u>**

For each PGS (**MDD anxiety, ADHD, EA**), we conducted a trio approach with generalized linear mixed models, to account for individual level (across time) and family level variation (siblings), to estimate the direct and indirect genetic effects on change in depressive symptoms at age 8 and 14. Although the result of model a and b below showed extremely large d.f which we didn't interpret the result further.

a. *Trio model to investigate genetic effects on depression across age 8 and 14*
depression ~ child_PGS + mother_PGS + father_ PGS + age + (1|child_ID) + (1|family_ID)

b. *Adjusting for covariates*

depression ~ child_PGS + mother _PGS + father_PGS + age+ parity+ sex+ HHincome+birth_year+ maternal_age+paternal_age +mother_edu+ father_edu + (1|child_ID) + (1|family_ID)

c. *Incorporate interaction terms to test genetic effects on emerging depressive symptoms (relevant terms in bold typeface)*

depression ~ child_PGS + mother_PGS + father_PGS + age + parity + sex + HHincome + mother_edu + father_edu + birth_year + maternal_age + paternal_age + +**child_PGS*age** +**mother_PGS*age** + **father_PGS*age** + (1|child_ID) + (1|family_ID)

NB – in model C, terms for ***interactions between age/waves and covariates*** were also included which is not depicted in c for brevity.

The interaction term between time and the child PGS capture the direct genetic effect on the emerging variation depressive symptoms at age 14 compared to age 8, while the interactions between time and, respectively, mother and father PGS capture the effect of indirect genetic effects on emerging variation depressive symptoms. The same model applied separately to each PGS.

#### Sensitivity analysis

Complete case analysis was used as a sensitivity to compare estimates with imputed analysis results. Sensitivity analyses using complete case data (CCA) were conducted to assess any impact of multiple imputation on the results beyond the expected increase in precision. Differences in the direction of associations or in statistical significance between CCA and multiple imputation (MI) analyses would indicate that the imputation procedure has materially influenced the estimates.

**Model building**

The appropriateness of the model was checked starting from intercept only model to the complex model with interactions and random intercepts. Based on the LRT it is confirmed that including both individual level and family level random intercepts were necessarily with p-value <0.001.

**Multicollinearity check**

We calculated variance inflation factors (VIFs) to assess multicollinearity among predictors (21) in both the CCA and MMI models. Because VIFs cannot be obtained from mixed-effects models, we computed without random effects. All predictors had VIFs below 4, indicating no problematic multicollinearity among the covariates.

#### Result 1

The availability of a well-powered, genotyped trio sample in MoBa strengthens the reliability of our estimates and enhances our ability to detect genetic effects with precision. Recognising that population-based cohort studies, including MoBa, are often affected by attrition we applied multilevel multiple imputation (MMI) under the assumption of missing at random (MAR), specifically addressing item-level missingness. Differences in results between CCA and MMI, where analyses using CCA is known to be biased under MAR, indicate that our use of MMI was an important strength of the study.

Although direct genetic effect estimates from polygenic scores adjusted for parental genotypes are less prone to bias than unadjusted estimates, they remain observational. Nonetheless, the removal of bias associated with parental genotypes in these models brings the estimates of direct genetic effects closer to causal estimates than would be possible in a population-based design (22).

Table S9- A trio analysis without adjusting for covariates: Assessing association of genetic liabilities and children's depressive symptoms at age 8 – on imputed data

| Family member | PGS | β | **Before FDR *p* value** | **FDR *p* value** | **95% CI** |
|---|---|---|---|---|---|
| Child | MDD | 0.022 | 0.01129 | 0.0226 | 0.0051,0.0393 |
| | ADHD | 0.024 | 0.004 | 0.0160 | 0.008, 0.041 |
| | Anxiety | 0.01 | 0.259 | 0.2590 | -0.007, 0.027 |
| | EA | -0.019 | 0.042 | 0.056 | -0.037, -0.001 |
| | Mothers trio estimates | | | | |
| Mothers | MDD | 0.016 | 0.04141 | 0.0552 | 0.0007,0.0321 |
| | ADHD | 0.01 | 0.192 | 0.2920 | -0.005, 0.041 |
| | Anxiety | 0.016 | 0.036 | 0.0552 | 0.001, 0.031 |
| | EA | 0.033 | 1.224e-05 | 0.00005 | 0.019, 0.048 |
| | Fathers trio estimate | | | | |
| Fathers | MDD | 0.003 | 0.66773 | 0.8903 | -0.0115, 0.0179 |
| | ADHD | 0.004 | 0.611 | 0.8903 | -0.011, 0.018 |
| | Anxiety | 0.0050 | 0.103 | 0.4120 | -0.002, 0.027 |
| | EA | -0.001 | 0.923 | 0.9230 | -0.015, 0.014 |

**Note:** MDD- major depressive disorder, ADHD-Attention Deficient/Hyperactivity disorder, EA-Educational attainment.

Table S10- A trio analysis without adjusting for covariates: Assessing association of genetic liabilities and children's depressive symptoms at age 14 – on imputed data

| Family member | PGS | β | **Before FDR *p* value** | **FDR *p* value** | **95% CI** |
|---|---|---|---|---|---|
| Child | MDD | 0.06285 | 7.714e-10 | 3.086e-09 | 0.0435, 0.0822 |
| | ADHD | 0.032 | 0.001 | 0.002 | 0.013, 0.050 |
| | Anxiety | 0.024 | 0.010 | 0.013 | 0.006, 0.042 |
| | EA | -0.017 | 0.097 | 0.097 | -0.037,0.003 |
| | Mothers trio estimates | | | | |
| Mothers | MDD | 0.012 | 0.1533 | 0.204 | -0.005, 0.029 |
| | ADHD | 0.016 | 0.065 | 0.204 | -0.001, 0.033 |
| | Anxiety | 0.013 | 0.139 | 0.204 | -0.004,0.030 |
| | EA | 0.003 | 0.776 | 0.776 | -0.015,0.020 |
| | Fathers trio estimate | | | | |
| Fathers | MDD | 0.012 | 0.129 | 0.348 | -0.003, 0.027 |
| | ADHD | 0.003 | 0.715 | 0.715 | -0.012, 0.018 |
| | Anxiety | 0.011 | 0.174 | 0.348 | -0.005,0.026 |
| | EA | -0.005 | 0.530 | 0.706 | -0.021,0.011 |

Table S11: Associations between genetic liabilities and children's depressive symptoms at age 8 and 14 *without* adjustment for parental PGS and control for measured confounders (Bivariate analysis).

| **Wave of measurement** | **Family member** | **PGS** | **β** | **95% CI** | **FDR *p* value** |
|---|---|---|---|---|---|
| 8 years | Child | MDD | 0.032 | 0.021- 0.043 | <0.001 |
| | | ADHD | 0.031 | 0.019, 0.043 | <0.001 |
| | | Anxiety | 0.024 | 0.012, 0.036 | 0.86 |
| | | EA | -0.001 | -0.014, 0.011 | 0.86 |
| | Mother | MDD | 0.028 | 0.016, 0.040 | <0.001 |
| | | ADHD | 0.023 | 0.010,0.035 | <0.001 |
| | | Anxiety | 0.021 | 0.009, 0.034 | <0.001 |
| | | EA | 0.023 | 0.010, 0.035 | <0.001 |
| | Father | MDD | 0.015 | 0.003, 0.027 | 0.020 |
| | | ADHD | 0.017 | 0.005,0.028 | 0.013 |
| | | Anxiety | 0.017 | 0.005,0.029 | 0.013 |
| | | EA | -0.007 | -0.019, 0.005 | 0.263 |
| 14 years | Child | MDD | 0.075 | 0.061, 0.089 | <0.001 |
| | | ADHD | 0.041 | 0.027, 0.055 | <0.001 |
| | | Anxiety | 0.036 | 0.022, 0.049 | <0.001 |
| | | EA | -0.019 | -0.033, -0.004 | 0.014 |
| | Mother | MDD | 0.044 | 0.030, 0.059 | <0.001 |
| | | ADHD | 0.032 | 0.018, 0.047 | <0.001 |
| | | Anxiety | 0.025 | 0.011, 0.039 | <0.001 |
| | | EA | -0.008 | -0.023, 0.007 | 0.303 |
| | Father | MDD | 0.044 | 0.031, 0.056 | 0.044 |
| | | ADHD | 0.020 | 0.007, 0.032 | 0.005 |
| | | Anxiety | 0.023 | 0.010, 0.036 | 0.003 |
| | | EA | -0.014 | -0.028, -0.001 | 0.042 |

**Note:** MDD- major depressive disorder, ADHD-Attention Deficient/Hyperactivity disorder, EA-Educational attainment.

Table S12: Association of genetic liabilities and children's depressive symptoms at age 8 and 14 in *fully adjusted* trio PGS models

| **Wave of measurement** | **Family member** | **PGS** | **β** | **95% CI** | **FDR *p* value** |
|---|---|---|---|---|---|
| 8 years | Child | MDD | 0.023 | 0.006, 0.040 | 0.018 |
| | | ADHD | 0.024 | 0.008, 0.041 | 0.016 |
| | | Anxiety | 0.009 | -0.008, 0.026 | 0.315 |
| | | EA | -0.020 | -0.038, -0.002 | 0.045 |
| | Mother | MDD | 0.016 | 0.000, 0.032 | 0.0626 |
| | | ADHD | 0.010 | -0.005, 0.025 | 0.199 |
| | | Anxiety | 0.017 | 0.001, 0.032 | 0.062 |
| | | EA | 0.040 | 0.024,0.055 | <0.001 |
| | Father | MDD | 0.001 | -0.014, 0.016 | 0.893 |
| | | ADHD | 0.002 | -0.013, 0.016 | 0.893 |
| | | Anxiety | 0.028 | -0.004, 0.026 | 0.464 |
| | | EA | 0.009 | -0.006, 0.024 | 0.464 |
| 14 years | Child | MDD | 0.063 | 0.044,0.083 | <0.001 |
| | | ADHD | 0.030 | 0.012, 0.049 | 0.002 |
| | | Anxiety | 0.023 | 0.005, 0.041 | 0.0187 |
| | | EA | -0.018 | -0.038, 0.002 | 0.083 |
| | Mother | MDD | 0.010 | -0.007,0.027 | 0.292 |
| | | ADHD | 0.013 | -0.003, 0.030 | 0.292 |
| | | Anxiety | 0.012 | -0.005, 0.029 | 0.292 |
| | | EA | 0.010 | -0.008,0.028 | 0.292 |
| | Father | MDD | 0.010 | -0.005,0.025 | 0.476 |
| | | ADHD | 0.0005 | -0.014, 0.015 | 0.951 |
| | | Anxiety | 0.009 | -0.006, 0.025 | 0.476 |
| | | EA | 0.001 | -0.015, 0.018 | 0.951 |

Table S13: Association of genetic liability on variation in symptoms of depression in early adolescence

| PGS | Family member | β | 95% CI | FDR *p* value |
|---|---|---|---|---|
| MDD:age | Child | 0.041 | 0.017, 0.065 | 0.004 |
| | Mother | -0.006 | -0.028, 0.016 | 0.770 |
| | Father | 0.008 | -0.012, 0.029 | 0.894 |
| ADHD:age | Child | 0.006 | -0.018, 0.029 | 0.825 |
| | Mother | 0.003 | -0.019, 0.025 | 0.771 |
| | Father | -0.002 | -0.022, 0.018 | 0.895 |
| Anx:age | Child | 0.014 | -0.010, 0.037 | 0.508 |
| | Mother | 0.017 | 0.001, 0.032 | 0.061 |
| | Father | -0.002 | -0.022, 0.018 | 0.895 |
| EA:age | Child | 0.003 | -0.022, 0.027 | 0.838 |
| | Mother | -0.029 | -0.053, -0.007 | 0.050 |
| | Father | -0.007 | -0.028, 0.013 | 0.894 |
| MDD | Child | 0.016 | 0.006, 0.039 | 0.017 |
| | Mother | 0.016 | 0.00037, 0.03185 | 0.060 |
| | Father | 0.00099 | -0.014, 0.016 | 0.895 |
| ADHD | Child | 0.024 | 0.008, 0.041 | 0.016 |
| | Mother | 0.009 | -0.005, 0.025 | 0.196 |
| | Father | 0.002 | -0.013, 0.016 | 0.895 |
| Anxiety | Child | 0.009 | -0.008, 0.026 | 0.317 |
| | Mother | 0.017 | 0.001, 0.032 | 0.061 |
| | Father | 0.011 | -0.004, 0.026 | 0.470 |
| EA | Child | -0.019 | -0.038, -0.002 | 0.042 |
| | Mother | 0.039 | 0.024, 0.055 | <0.001 |
| | Father | 0.009 | -0.006, 0.024 | 0.470 |

**Sex stratified associations between genetic liabilities to MDD ADHD, Anxiety and EA and variation in early adolescence depressive symptoms in a fully adjusted Trio PGS model**

The model-predicted the variation in early adolescence depressive symptoms from age 8 and 14 based on childrens's MDD, ADHD, Anxiety and EA genetic liability are shown, stratified by sex (Fig S4).

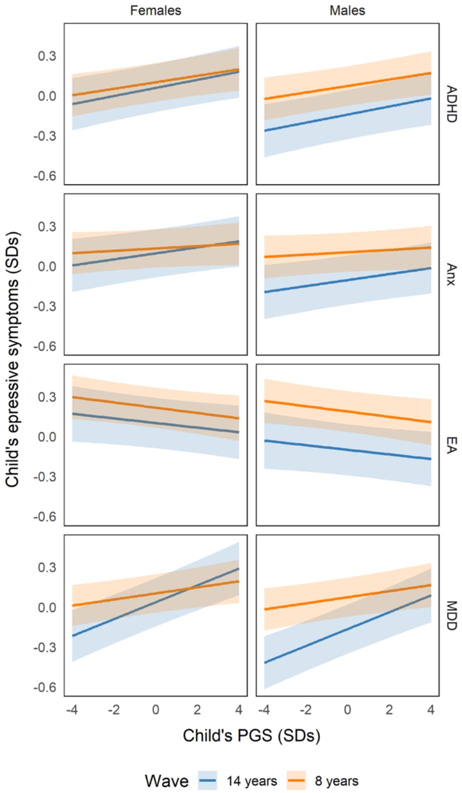

***Figure S4.*** *Sex-stratified relationships between children's own genetic liabilities to MDD,ADHD, Anxiety and EA and their depressive symptoms, as predicted by the main effects and interactions of MDD, ADHD, Anxiety and EA PGS and age at measurement in trio-PGS models, with other factors held constant*

The model-predicted the variation in early adolescence depressive symptoms from age 8 and 14 based on mother's MDD, ADHD, Anxiety and EA genetic liability are shown, stratified by sex (Fig S5).

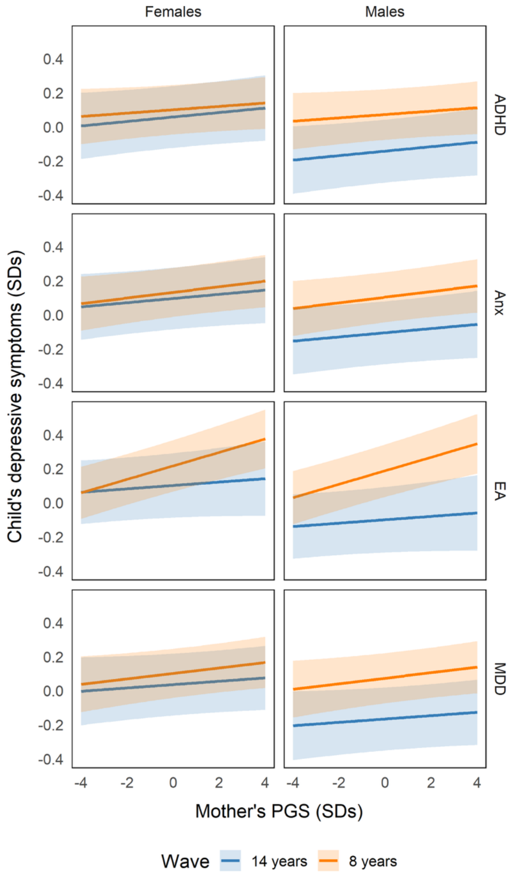

***FigureS5.*** *Sex-stratified relationships between mothers' genetic liabilities to MDD, ADHD, anxiety, EA and children depressive symptoms, as predicted by the main effects and interactions of MDD MDD, ADHD, anxiety, EA PGS and age at measurement in trio-PGS models, with other factors held constant*

The model-predicted the variation in early adolescence depressive symptoms from age 8 and 14 based on father's MDD, ADHD, Anxiety and EA genetic liability are shown, stratified by sex (Fig S6).

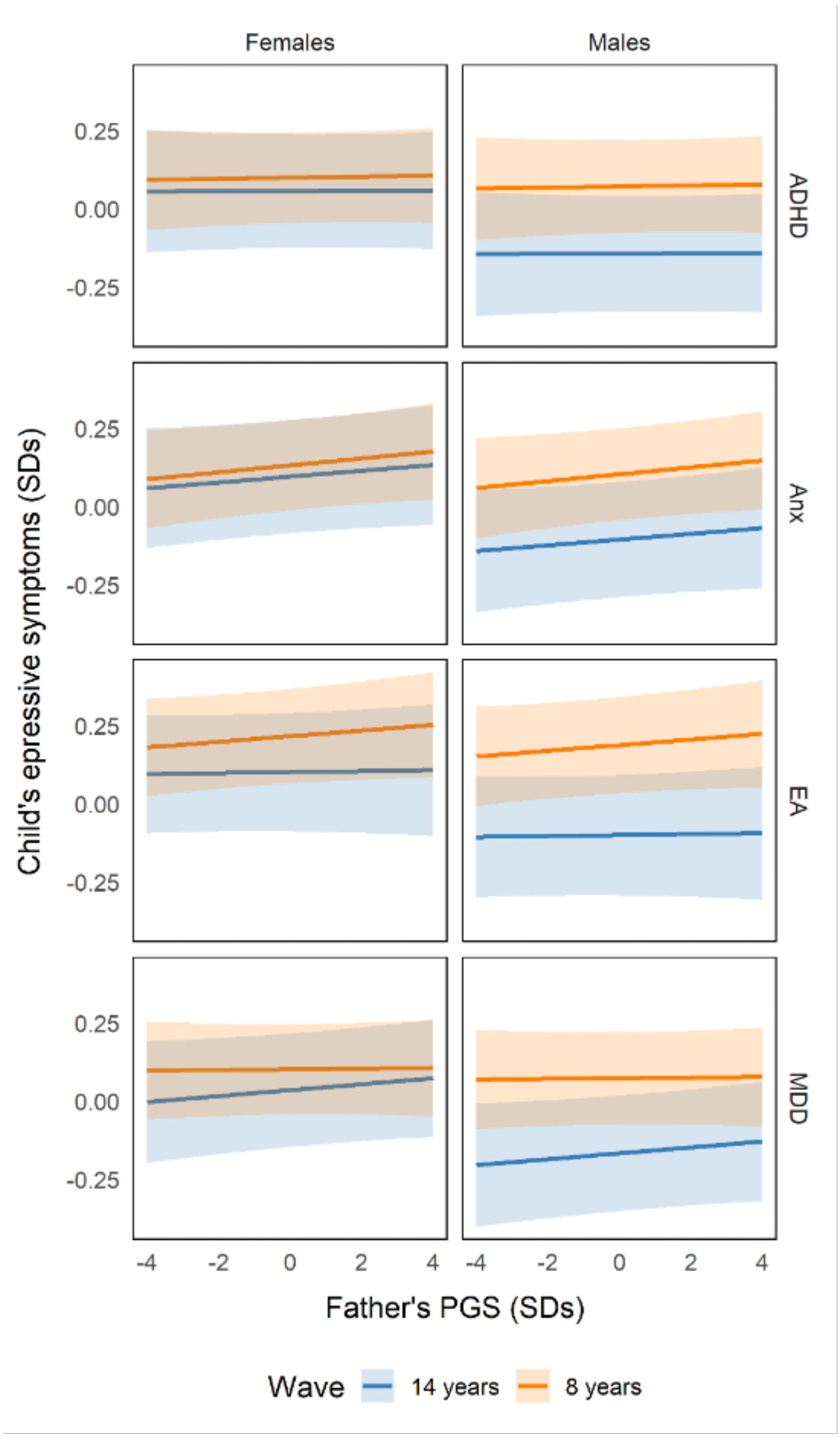

***FigureS6:*** *Sex-stratified relationships between fathers' genetic liabilities to MDD ADHD Anxiety and EA and children's depressive symptoms, as predicted by the main effects and interactions of MDD ADHD Anxiety and EA PGS and age at measurement in trio-PGS models, with other factors held constant*

#### Result 2

**Complete case analysis results**

**CCA results age 8**: a bivariate analysis from CCA indicated that except children's genetic liability to EA, all other exposures MDD, ADHD and Anxiety PGS found to have significant association with their own depressive symptoms at age 8. Whereas, adjusting for parental genetic effect (trio analysis) children EA PGS becomes significant ((β=-0.03, 95% CI: -0.05 – -0.004, P=0.024) while child Anxiety PGS becomes insignificant. However, the estimates on a trio analysis for MDD and ADHD were attenuated but remained significant. This trio results remain the same even after adjusting for SES covariates.

On the other hand, on a bivariate analysis, genetic liability of both parents to all exposures were found significantly associated with children's depressive symptoms at age 8, except fathers' genetic liability to EA. However, on a trio analysis fathers' genetic liability to all exposure becomes statistically non-significant, while *maternal genetic liabilities* to all exposure *remain significant*. These trio results remain the same after adjusting to SES, except, mothers' genetic liability to *ADHD becomes non-significant*.

Table S14. A CCA trio estimate of direct and indirect genetic effect on depressive symptoms at age 8 adjusted for SES.

| | β | C.I | P-value FDR |
|---|---|---|---|
| Children CCA trio estimates | | | |
| MDD | 0.04 | 0.02 – 0.06 | 0.001 |
| ADHD | 0.04 | 0.02 – 0.06 | 0.001 |
| Anx | 0.01 | -0.01 – 0.03 | 0.280 |
| EA | -0.03 | -0.05 – -0.01 | 0.006 |
| Mothers CCA trio estimates | | | |
| MDD | 0.03 | 0.01 – 0.05 | 0.003 |
| ADHD | 0.02 | -0.00 – 0.03 | 0.079 |
| Anx | 0.03 | 0.01 – 0.05 | 0.003 |
| EA | 0.06 | 0.04 – 0.07 | <0.001 |
| Fathers CCA trio estimates | | | |
| MDD | -0.0009 | -0.02 – 0.02 | 0.922 |
| ADHD | 0.002 | -0.02 – 0.02 | 0.922 |
| Anx | 0.02 | 0.00 – 0.04 | 0.069 |
| EA | 0.02 | 0.00 – 0.04 | 0.075 |

Table S15. A CCA trio estimate of direct and indirect genetic effect on depressive symptoms at age 14 adjusted for SES.

| Role | PGS | β | C.I | P-value FDR |
|---|---|---|---|---|
| Childs | MDD | 0.18 | 0.14 – 0.23 | <0.001 |
| | ADHD | 0.05 | 0.02 – 0.08 | <0.001 |
| | Anx | 0.03 | 0.00 – 0.05 | <0.001 |
| | EA | -0.03 | -0.06 – -0.00 | 0.037 |
| Mothers | MDD | 0.03 | -0.01 – 0.08 | 0.105 |
| | ADHD | 0.02 | -0.00 – 0.04 | 0.105 |
| | Anx | 0.02 | 0.00 – 0.05 | 0.105 |
| | EA | 0.02 | -0.00 – 0.04 | 0.105 |
| Fathers | MDD | 0.05 | 0.01 – 0.10 | 0.024 |
| | ADHD | 0.01 | -0.02 – 0.03 | 0.798 |
| | Anx | 0.03 | 0.01 – 0.05 | 0.024 |
| | EA | 5.804e-04 | -0.02 – 0.02 | 0.962 |

**CCA results age 14**: On a bivariate and trio complete case analysis, children genetic liability to all exposure showed evidence of association with their own depressive symptoms at age 14. This result remains the same after adjusting for SES-s*uggesting direct genetic effect at age 14.*

On a bivariate analysis, mothers and fathers' genetic liability to all exposure were significantly associated with adolescence depressive symptoms except for maternal EA. To the contrary, on a trio analysis we observe that *maternal genetic liability* to all of the exposures showed *no association* with children's depressive symptoms at age 14. However, *fathers' genetic liability to MDD* (($\beta$ =0.04, 95% CI: 0.01 – 0.06, P=0.009) and *anxiety* (($\beta$ =0.03, 95% C.I: 0.01 – 0.05, P=0.024) showed association. These trio results remain the same even after adjusted for SES--indicating that there is indirect genetic effect from the father but not from the mother at age 14.

Estimating emerging variation in depressive symptoms on a CCA showed that children genetic liability to ADHD and EA shown evidence of association with their own depressive symptoms that was stable *across ages (8 and 14)* while child genetic liability to MDD (($\beta_{PGS*wave}$=0.06, 95% CI: 0.03 – 0.09, P<0.001) was the only exposure showed age-specific depressive symptoms indicating direct genetic liability. A one s.d increase in children MDD PGS will increase their own depressive symptoms by 6% SD at age 14 compared to at age 8.

Fathers' genetic liability to all exposures showed limited evidence of association with children's depressive symptoms. To the contrary to the imputed results, maternal genetic liability to MDD (($\beta$=0.03, 95% CI: 0.01 – 0.05, P=0.002), Anxiety (($\beta$=0.03, 95% CI: 0.01 – 0.05, P=0.003) and EA ($\beta$= 0.05, 95% C.I=0.03-0.07, P<0.001) showed strong association with depressive symptoms across age 8 and 14 (stable effect) . There was no age specific effect from the parents genetic liabilities in the CCA.

Table S16: CCA on variation in depressive symptoms from age 8 to age 14 adjusted for covariates.

| **Role** | **PGS** | β | **C.I** | **P-value FDR** |
|---|---|---|---|---|
| **Child** | MDD | 0.03 | 0.01 – 0.05 | 0.002 |
| | ADHD | 0.04 | 0.02 – 0.06 | 0.0006 |
| | Anx | 0.01 | -0.01 – 0.03 | 0.294 |
| | EA | -0.03 | -0.05 – -0.01 | 0.011 |
| | MDD: AGE | 0.06 | 0.03 – 0.09 | 0.0002 |
| | ADHD:AGE | 0.02 | -0.01 – 0.05 | 0.417 |
| | Anx:AGE | 0.02 | -0.01 – 0.04 | 0.417 |
| | EA:AGE | -4.632e-03 | -0.04 – 0.03 | 0.769 |
| **Mothers** | MDD | 0.03 | 0.01 – 0.05 | 0.002 |
| | ADHD | 0.01 | -0.00 – 0.03 | 0.117 |
| | Anx | 0.03 | 0.01 – 0.05 | 0.003 |
| | EA | 0.05 | 0.04 – 0.07 | <0.001 |
| | MDD:Age | -0.01 | -0.04 – 0.02 | 0.747 |
| | ADHD:Age | 0.01 | -0.02 – 0.03 | 0.747 |
| | Anx:Age | **-3.457e-03** | -0.03 – 0.02 | 0.792 |
| | EA:Age | -0.03 | -0.06 – -0.01 | 0.060 |
| **Fathers** | MDD | -4.948e-04 | -0.02 – 0.02 | 0.956 |
| | ADHD | **1.002e-03** | -0.02 – 0.02 | 0.956 |
| | Anx | 0.02 | 0.00 – 0.04 | 0.064 |
| | EA | 0.02 | 0.00 – 0.04 | 0.084 |
| | MDD:Age | 0.03 | 0.01 – 0.06 | 0.064 |
| | ADHD:Age | 3.081e-03 | -0.02 – 0.03 | 0.812 |
| | Anx:Age | 0.01 | -0.02 – 0.03 | 0.691 |
| | EA:Age | -0.01 | -0.04 – 0.01 | 0.536 |

**<u>Result 3</u>**

**Comparison of MMI and CCA**

The CCA direct genetic effect estimates aligned with the imputed data results at age 8 and 14. At age 8, the CCA results showed strong evidence of maternal indirect genetic effect association with depressive symptoms. This result was contrary to imputed data results.

At age 14, we have seen that there were indirect genetic effects from the father but not from the mother at age 14 in the CCA. These results indicated similar trend with the imputed data on the effect of indirect genetic effect from the mother, but not from paternal genetic effect at age 14. In the longitudinal model to estimate variation in emerging depressive symptoms, direct genetic effect estimates and indirect genetic effect from the fathers on a CCA align with the imputed data. Whereas maternal genetic effect estimates on a CCA showed significant difference with the imputed data results.

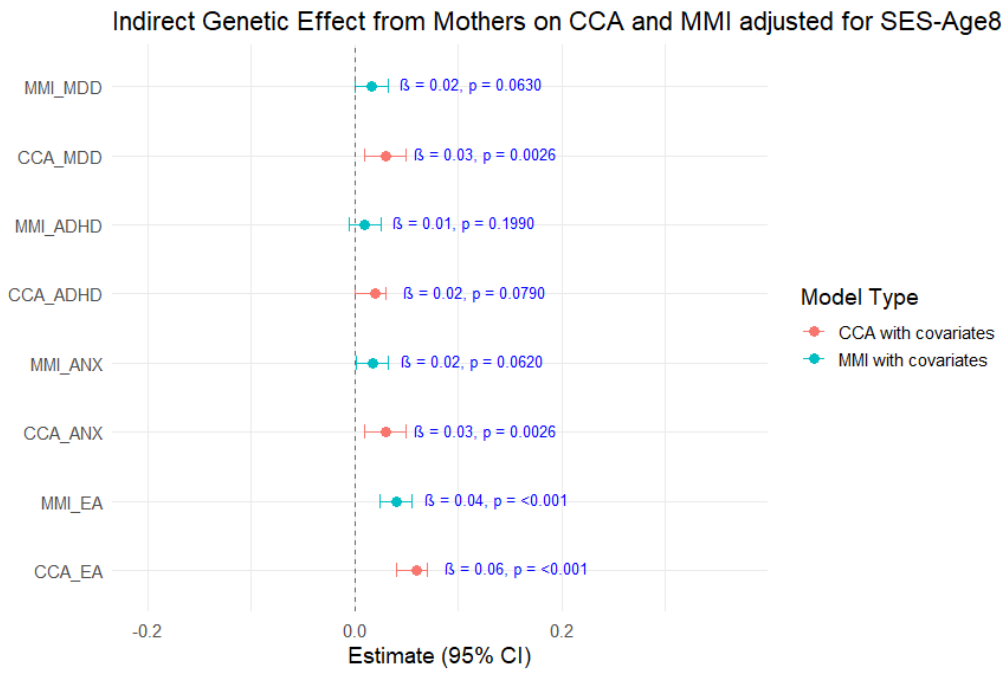

Fig S6- CCA vs Imputed Mothers genetic effect estimates on children depressive symptoms at age 8 adjusted for confounders

#### Other sensitivity analysis

Using mother reported full 13-item scale on an imputed data, depressive symptoms at age 8 showed associations with children's own genetic liabilities for MDD, ADHD, and EA , similar to the 6-item measure. Maternal genetic liabilities for MDD and EA showed indirect genetic effects, whereas no paternal indirect effects were observed for any of the genetic liabilities at age 8 (Supplementary Table S17). These parental effects showed similar trends compared to the 6-item measure except for maternal MDD, which showed little evidence of an association in the 6-item analysis.

Based on CCA with child-reported items at age 14 (since the MMI model failed to converge) children's genetic liabilities for MDD, ADHD and anxiety showed evidence of association with depressive symptoms, with little evidence of parental genetic effects. These results aligned with the mother-reported 6-item version based on imputed data (S18).

Table S17: A trio model adjusted with covariates based on mother reported *13 items* at age 8.

| Role | | β | SE | C.I | FDR P-value |
|---|---|---|---|---|---|
| **Child PGS** | MDD | 0.026 | 0.009 | 0.009, 0.043 | 0.004 |
| | ADHD | 0.042 | 0.008 | 0.025, 0.058 | <0.001 |
| | Anxiety | 0.012 | 0.008 | -0.004, 0.028 | 0.130 |
| | EA | -0.036 | 0.009 | -0.053, -0.018 | <0.001 |
| **Mothers PGS** | MDD | 0.022 | 0.008 | 0.006, 0.038 | 0.016 |
| | ADHD | 0.008 | 0.007 | -0.006, 0.023 | 0.261 |
| | Anxiety | 0.014 | 0.007 | -0.001, 0.029 | 0.080 |
| | EA | 0.043 | 0.008 | 0.027, 0.060 | <0.001 |
| **Fathers PGS** | MDD | -0.001 | 0.007 | -0.015, 0.014 | 0.921 |
| | ADHD | -0.002 | 0.007 | -0.015, 0.012 | 0.921 |
| | Anxiety | 0.010 | 0.007 | -0.004, 0.024 | 0.364 |
| | EA | 0.015 | 0.007 | 7.67e-05, 0.029 | 0.196 |

Table S18: A trio CCA model adjusted with covariates based on child reported *items* at age 14.

| Role | | Estimate | C.I | P-val before | P-value FDR |
|---|---|---|---|---|---|
| **Child PGS** | **MDD** | 0.65 | 0.50 – 0.80 | < 2e-16 | 8.0e-16 |
| | **ADHD** | 0.45 | 0.30 – 0.61 | 8.79e-09 *** | 1.7e-08 |
| | **Anx** | 0.26 | 0.11 – 0.41 | 0.001 | 1.3e-03 |
| | **EA** | -0.15 | -0.31 – 0.01 | 0.075 | 7.50e-02 |
| **Mothers PGS** | **MDD** | 0.10 | -0.03 – 0.23 | 0.143 | 0.286 |
| | **ADHD** | 0.08 | -0.06 – 0.21 | 0.260 | 0.3466 |
| | **Anx** | 0.06 | -0.07 – 0.19 | 0.375 | 0.37500 |
| | **EA** | 0.16 | 0.02 – 0.30 | 0.026 | 0.10400 |
| **Fathers PGS** | **MDD** | 0.08 | -0.05 – 0.22 | 0.223 | 0.29733 |
| | **ADHD** | -0.05 | -0.19 – 0.08 | 0.423 | 0.42300 |
| | **Anx** | 0.10 | -0.03 – 0.23 | 0.149 | 0.29733 |
| | **EA** | 0.15 | 0.01 – 0.29 | 0.036 | 0.14400 |

#### References

1. Angold, A., Costello, E. J., & Messer, S. C. (1996). Development of a short questionnaire for use in epidemiological studies of depression in children and adolescents. Vol. 5. 1996 Dec 1;5(4):237–49.

2. Choi SW, Mak TSH, O'Reilly PF. Tutorial: a guide to performing polygenic risk score analyses. Nat Protoc. 2020 Sep 1;15(9):2759–72. doi:10.1038/s41596-020-0353-1

3. Allegrini AG, Baldwin JR, Barkhuizen W, Pingault J. Research Review: A guide to computing and implementing polygenic scores in developmental research. Child Psychology Psychiatry. 2022 Oct;63(10):1111–24. doi:10.1111/jcpp.13611

4. Mostafavi H, Harpak A, Agarwal I, Conley D, Pritchard JK, Przeworski M. Variable prediction accuracy of polygenic scores within an ancestry group. eLife. 2020 Jan 30;9:e48376. doi:10.7554/eLife.48376

5. McIntosh AM, Lewis CM, Mark J Adams for the Psychiatric Genomics Consortium Major Depressive Disorder Working Group. Genome-wide study of half a million individuals with major depression identifies 697 independent associations, infers causal neuronal subtypes and biological targets for novel pharmacotherapies [Internet]. 2024 [cited 2024 Nov 27]. Available from: http://medrxiv.org/lookup/doi/10.1101/2024.04.29.24306535 doi:10.1101/2024.04.29.24306535

6. Demontis D, Walters GB, Athanasiadis G, Walters R, Therrien K, Nielsen TT, et al. Genome-wide analyses of ADHD identify 27 risk loci, refine the genetic architecture and implicate several cognitive domains. Nat Genet. 2023 Feb;55(2):198–208. doi:10.1038/s41588-022-01285-8

7. Strom NI, Verhulst B, Bacanu SA, Cheesman R, Purves KL, Gedik H, et al. Genome-wide association study of major anxiety disorders in 122,341 European-ancestry cases identifies 58 loci and highlights GABAergic signaling [Internet]. Genetic and Genomic Medicine; 2024 [cited 2025 Nov 10]. Available from: http://medrxiv.org/lookup/doi/10.1101/2024.07.03.24309466 doi:10.1101/2024.07.03.24309466

8. Okbay A, Wu Y, Wang N, Jayashankar H, Bennett M, Nehzati SM, et al. Polygenic prediction of educational attainment within and between families from genome-wide association analyses in 3 million individuals. Nat Genet. 2022 Apr;54(4):437–49. doi:10.1038/s41588-022-01016-z

9. Privé F, Arbel J, Vilhjálmsson BJ. LDpred2: better, faster, stronger. Schwartz R, editor. Bioinformatics. 2021 Apr 1;36(22–23):5424–31. doi:10.1093/bioinformatics/btaa1029

10. Corfield EC, Shadrin AA, Frei O, Rahman Z, Lin A, Athanasiu L, et al. The Norwegian Mother, Father, and Child cohort study (MoBa) genotyping data resource: MoBaPsychGen pipeline v.1 [Internet]. 2022 [cited 2025 Sep 11]. Available from: http://biorxiv.org/lookup/doi/10.1101/2022.06.23.496289 doi:10.1101/2022.06.23.496289

11. Privé F, Albiñana C, Arbel J, Pasaniuc B, Vilhjálmsson BJ. Inferring disease architecture and predictive ability with LDpred2-auto. The American Journal of Human Genetics. 2023 Dec;110(12):2042–55. doi:10.1016/j.ajhg.2023.10.010

12. Little RJA. A Test of Missing Completely at Random for Multivariate Data with Missing Values. Journal of the American Statistical Association. 1988 Dec;83(404):1198–202. doi:10.1080/01621459.1988.10478722

13. Rubin DB. Inference and missing data. Biometrika. 1976;63(3):581–92. doi:10.1093/biomet/63.3.581

14. Kontopantelis E, White IR, Sperrin M, Buchan I. Outcome-sensitive multiple imputation: a simulation study. BMC Med Res Methodol. 2017 Dec;17(1):2. doi:10.1186/s12874-016-0281-5

15. Van Ginkel JR, Linting M, Rippe RCA, Van Der Voort A. Rebutting Existing Misconceptions About Multiple Imputation as a Method for Handling Missing Data. Journal of Personality Assessment. 2020 May 3;102(3):297–308. doi:10.1080/00223891.2018.1530680

16. Mainzer RM, Nguyen CD, Carlin JB, Moreno-Betancur M, White IR, Lee KJ. A comparison of strategies for selecting auxiliary variables for multiple imputation. Biometrical J. 2024 Jan;66(1):2200291. doi:10.1002/bimj.202200291

17. Bartlett JW, Seaman SR, White IR, Carpenter JR, for the Alzheimer's Disease Neuroimaging Initiative*. Multiple imputation of covariates by fully conditional specification: Accommodating the substantive model. Stat Methods Med Res. 2015 Aug;24(4):462–87. doi:10.1177/0962280214521348

18. Rubin DB. Multiple Imputation for Nonresponse in Surveys [Internet]. 1st edn. Wiley; 1987 [cited 2026 Apr 3]. (Wiley Series in Probability and Statistics). Available from:

https://onlinelibrary.wiley.com/doi/book/10.1002/9780470316696 doi:10.1002/9780470316696

19. Van Buuren S. Flexible Imputation of Missing Data, Second Edition [Internet]. 2nd edn. Second edition. | Boca Raton, Florida : CRC Press, [2019] |: Chapman and Hall/CRC; 2018 [cited 2025 Sep 30]. Available from: https://www.taylorfrancis.com/books/9780429492259 doi:10.1201/9780429492259

20. White IR, Royston P, Wood AM. Multiple imputation using chained equations: Issues and guidance for practice. Statistics in Medicine. 2011 Feb 20;30(4):377–99. doi:10.1002/sim.4067

21. Draper NR, Smith H. Applied Regression Analysis [Internet]. 1st edn. Wiley; 1998 [cited 2026 Feb 20]. (Wiley Series in Probability and Statistics). Available from: https://onlinelibrary.wiley.com/doi/book/10.1002/9781118625590 doi:10.1002/9781118625590

22. Veller C, Coop GM. Interpreting population- and family-based genome-wide association studies in the presence of confounding. Moorjani P, editor. PLoS Biol. 2024 Apr 11;22(4):e3002511. doi:10.1371/journal.pbio.3002511
